# Glycoprotein Acetyls and Depression: testing for directionality and potential causality using longitudinal data and Mendelian randomization analyses

**DOI:** 10.1101/2022.12.06.22283149

**Authors:** Daisy C P Crick, Eleanor Sanderson, Hannah Jones, Neil Goulding, Maria Carolina Borges, Gemma Clayton, Alice R Carter, Sarah Halligan, Deborah A Lawlor, Golam M Khandaker, Abigail Fraser

## Abstract

**Background:** Inflammation is implicated in depression, but the issue of causality remains unclear.

**Objectives:** To investigate potential causality and direction of effect between inflammation and depression.

**Methods:** Using data from the ALSPAC birth cohort (n=4021), we used multivariable regression to investigate bidirectional longitudinal associations of GlycA and depression symptoms score and diagnosis, assessed at ages 18y and 24y.

We used two-sample Mendelian randomization (MR) to investigate potential causality and directionality. Genetic variants for GlycA were obtained from UK Biobank (UKBB) (N=115,078); for depression from the Psychiatric Genomics Consortium and UKBB (N=500,199); and for depressive symptoms (N=161,460) from the Social Science Genetic Association Consortium. In addition to the Inverse Variance Weighted (IVW) method, we used sensitivity analyses to strengthen causal inference. We conducted multivariable MR adjusting for body mass index (BMI) due to known genetic correlation between inflammation, depression and BMI.

**Results:** After adjusting for potential confounders we found no association between GlycA and depression symptoms score or *vice versa*. We observed an association between GlycA and depression diagnosis (OR=1.18, 95% CI: 1.03-1.36).

MR suggested no causal effect of GlycA on depression, but there was evidence of a causal effect of depression on GlycA (mean difference in GlycA = 0.09; 95% CI: 0.03-0.16), which was maintained in some, but not all, sensitivity analyses.

**Conclusion:** We found no consistent evidence for an effect of the inflammatory marker GlycA on depression. There was some evidence that depression may increase GlycA, but this may be confounded/mediated by BMI.

## Introduction

Major Depressive Disorder (MDD) is one of the most prevalent psychiatric disorders worldwide and a leading cause of disease burden ^(1)^ with a lifetime prevalence of 5-17%.^(2)^ The identification of causal risk factors for MDD could provide novel targets for both prevention and treatment.

An association between inflammation and depression has been identified in cross-sectional and meta-analyses of case/control studies. ^(3-5)^ However, in both study designs, the exposure and outcome are measured concurrently, meaning reverse causality cannot be ruled out. Comparatively, prospective cohort studies (which are better able to evaluate temporality/reverse causality) have found evidence of a bi-directional relationship between depression and inflammation.^(6-10)^ Although unmeasured confounding may still bias these results, further support for this bidirectional association has come from randomised control trials (RCTs) which are said to be the “gold standard” for epidemiological evidence. ^(11-16)^.

For example, an RCT found that an Interferon-alpha (IFN-α) based treatment for cancer raises serum-based cytokine levels which in turn predict the development of depressive symptoms.^(11)^ However, other factors could drive this association such as the treatment causing lower feelings of wellbeing and hence an increase in symptoms of depression.

Mendelian Randomisation (MR) ^(17,18)^ aims to estimate the causal effect of an exposure on an outcome, by utilising the random assortment of genetic variants from parents to offspring.^(19)^ MR has differing and unrelated sources of bias compared to multivariable regression analyses. Integrating results from different methods (triangulation) can enhance causal inference when results are concordant.^(20)^ A description of MR and its assumptions is provided in the supplement.

MR studies have begun to investigate the potential causal effect of inflammation on depressive symptoms.^(3,21)^ Ye et al., found genetically predicted higher CRP concentrations had a protective effect on depression and anxiety.^(21)^ In contrast, genetically predicted higher interleukin-6 (IL-6) levels increased risk of fatigue and sleep problems, but had no effect on overall depression score.^(3)^ Other MR studies have reported positive causal effects of soluble IL-6 receptor (sIL-6R) on depression ^(22)^ and IL-6 activity increased risk of depressive symptoms.^(21)^ However, there are a limited number of SNPs associated with circulating levels/concentrations of IL-6 at genome-wide significance that can be used as instrumental variables (IVs; N< 2).^(23)^ This makes interpretation of results difficult as the IL-6 IVS sometimes reflect soluble IL-6 receptors (e.g. ^(22)^) rather than circulating IL-6 levels.

Existing research has focused on the IL-6 cytokine and the acute-phase protein CRP (production of which is stimulated by IL-6 levels). Although, archetypal inflammatory markers, both IL-6 and CRP have been found to associate with anti-inflammatory roles within the immune system. ^(24)^ Additionally, both biomarkers show daily fluctuations ^(25,26)^ and respond to short-term stressors,^(27-32)^ suggesting that they are not ideal markers of chronic inflammation.^(33)^ Using biomarkers that better index chronic, systemic inflammation may help understand the directionality and mechanisms underlying the association between depression and inflammation.

Glycoprotein Acetyls (GlycA) is a nuclear magnetic resonance (NMR) biomarker which summarises the signals originating from glycan groups of certain acute-phase glycoproteins. GlycA becomes elevated in response to insults such as infection and physical injury.^(34)^ Further, it is elevated in patients with a diverse range of inflammation-linked chronic conditions ^(35,36)^ and due to its composite nature ^(34)^ displays long-term stability.^(37)^ Therefore, unlike CRP or IL-6, GlycA provides an overall index of inflammation in the body ^(38,39)^ and likely to be a superior marker of chronic systemic inflammation.^(34,40)^

Here we aim to understand the relationship between chronic systemic inflammation measured by GlycA and depression diagnosis/symptoms and vice versa. We investigate longitudinal associations using data from a population-based prospective UK birth cohort ^(41-43)^ and the potential causal effect using bidirectional MR.

## Methods

The Avon Longitudinal Study of Parents and Children (ALSPAC) birth cohort recruited a total of 14,541 pregnant women residing in the former county of Avon; in South-West England ^(41-43)^. Women were recruited if they had an expected delivery date between 1st April 1991 and 31st December 1992. When the oldest children were approximately 7 years of age, an attempt was made to bolster the initial sample with eligible cases who did not originally join the study. The total sample size for analyses using any data collected after the age of seven is therefore 15,454 pregnancies, resulting in 15,589 foetuses. Of these, 14,901 were alive at 1 year of age. The offspring, their mothers and the mother’s partners are regularly followed up. Additional information on ALSPAC is presented in the supplementary file.

To be eligible for inclusion in the current study, participants had to have one measure of GlycA and a completed Short Moods and Feelings Questionnaire (SMFQ) at 18y or 24y (N=4021). The participant flowchart is presented in supplementary Figure 1.

We used the SMFQ ^(44-47)^ administered at mean ages 18y and 24y in ALSPAC. The SMFQ comprises 13 items relating to depressive symptoms experienced in the last two weeks. Each item is rated on a 3-point scale (0=not true, 1=sometimes true, 2=true), giving a total score of 0 to 26. Higher scores are suggestive of a greater burden of depressive symptoms.

We also used the Clinical Interview Schedule-Revised (CIS-R) as a secondary measure. CIS-R is used to diagnose depression according to ICD-10 codes.^(48)^ A description of how the variable was created and why it was used is provided in the supplement. Given that the SMFQ demonstrates good discriminatory abilities for identification of depression as measured by the CIS-R ^(49)^ and the limited statistical power of the CIS-R binary measure, the SMFQ score was used as a primary outcome.

Plasma GlycA was quantified as part of a high-throughput proton (1H) Nuclear Magnetic Resonance (NMR) metabolomic trait platform at mean ages 18y and 24y.^(38)^ Details regarding sample/data processing and NMR analysis have been provided in the supplement and elsewhere.^(50,51)^

Maternal highest education qualification self-reported at 8-42 weeks gestation (degree/no degree) proxied childhood socio-economic position (SEP). Participants’ age (months), smoking status and drinking status (non-smoker, infrequent smoker and frequent smoker) and BMI were reported at ages 18y and 24y. Participants’ ethnicity was recorded and coded as white or non-white. These confounders were determined *a priori* based on known or plausible causes of both systemic inflammation and depression. See supplement for further information on the choice of primary measure and how confounder variables were created.

### Statistical analysis

Multivariable linear regression examined prospective associations (effect of exposure at 18y on outcome at 24y) between SMFQ scores and GlycA levels and vice versa. Results were presented sex combined (all p-values for sex interaction > 0.822). GlycA levels and SMFQ scores were logged and z-scored to allow comparison between both directions.

We used multiple imputation (MI) to impute missing exposure, outcome and confounder data in the eligible sample, creating 25 datasets with 250 iterations. This aimed to balance computational efficiency while ensuring consistency of results.^(52)^ The main and auxiliary variables are listed in supplementary table 1. Estimates were combined using Rubin’s rules.^(52-54)^

### Univariable bidirectional two-sample Mendelian Randomisation

We used summary data from European population GWAS of GlycA,^(55)^ MDD ^(56)^ and depressive symptom.^(57)^ The GlycA GWAS was undertaken in UK Biobank data (UKBB), the MDD GWAS was from the Psychiatric Genomics Consortium (PGC) and the depressive symptoms GWAS from the Social Science Genetic Association Consortium (SSGAC).

#### Instrument Selection

Genome Wide significant SNPs (P< 5.0 x10-8) were selected from the GWAS described above as instrumental variables for GlycA, MDD and depressive symptoms. The datasets were harmonised, aligning the genetic association for exposure and outcome on the effect allele using the effect allele frequency. Palindromic SNPs with allele frequencies of above 0.42 were considered strand ambiguous and removed. SNPs that were unavailable in the outcome datasets were replaced by proxy SNPs at R^2^>0.80. SNPs that had a minor allele frequency of 0.01 or less were excluded.

Following harmonization, there were 50 SNPs in the analysis having GlycA as the exposure and MDD as the outcome (hereafter GlycA-MDD analysis), 40 SNPs in the analysis having GlycA as the exposure and depressive symptoms as the outcome (hereafter GlycA-Depressive Symptoms analysis) and 47 SNPs in the analysis having MDD as an exposure and GlycA as the outcome (hereafter MDD-GlycA analysis). The differences in number of SNPs available for each analysis is because not all SNPs were available in each GWAS. Full details of the instrument selection process are presented in the supplementary materials. GWAS of depressive symptoms have found only two genome-wide significant SNPs and we therefore did not use depressive symptoms as an exposure. Harmonized genetic association data for selected SNPs are presented in supplementary tables 2-4 respectively.

#### Main analyses

The inverse variance weighted method (IVW method) ^(58)^ was used to estimate effects in the primary analysis and assumes there is no directional horizontal pleiotropy. MR-Egger, weighted median and weighted mode methods were used as sensitivity analyses.

Consistency in results between methods strengthens causal inference because they make different assumptions about pleiotropy, divergent results suggest a violation of the third MR assumption.^(59-61)^ We reported F-statistics as a measure of instrument strength and investigated MR assumptions using: Cochran’s Q, MR-Egger intercept, Radial MR and MR Pleiotropy Residual Sum and Outlier global test (MR-PRESSO). Results are presented post-Steiger filtering. See supplementary table 5 for a description of the MR methods and sensitivity analyses used.

#### Further Analyses

MR-Lap is a method that corrects for sample overlap, weak instrument bias and winner’s curse.^(62)^ It is used when the exposure and outcome are continuous and so was used for the GlycA-depressive symptoms analysis only. A description of MR-Lap is given in the supplement.

We replicated analyses using IL-6 as the inflammatory biomarker. We aimed to replicate previous null findings for IL-6 in relation to depressive symptoms and MDD ^(3,7,63-66)^ as a quality assurance test of the methodology and data sources that we used. The data sources, SNP selection and method of analysis is described in the supplement.

### Multivariable Mendelian Randomisation

Multivariable Mendelian Randomization (MVMR) is an extension of MR that estimates the direct causal effect of multiple exposures on an outcome^(67)^ and can be used to account for pleiotropic pathways. Due to the proinflammatory role of adipose tissue ^(68)^ and evidence of its bidirectional association with depression ^(69)^, we estimated the causal effects of depression on inflammation, independently of BMI. A detailed description of MVMR can be found in the supplement. We used the BMI GWAS with the largest number of SNPs (https://gwas.mrcieu.ac.uk/; data code: ukb-a-248).

#### Instrument selection

We used the same thresholds for genome-wide significance, LD and palindromic SNPs as in the univariable MR. The MDD SNPs and BMI SNPs were combined and harmonised with the GlycA data. There were 316 BMI SNPs after pruning and when combined with the MDD SNPs there was a final exposure instrument of 618 SNPs for the depression-GlycA MVMR. After harmonization with GlycA, 299 SNPs were available for the MVMR analysis. Full details of the SNP selection process are presented in the supplementary material. We only ran an MVMR for MDD and BMI as exposures in relation to the outcome GlycA because we found no evidence of an effect in the GlycA-MDD/GlycA-depressive symptoms analyses in the univariable MR.

We repeated the MVMR using only the genome-wide associated MDD SNPs used in our univariable MDD-GlycA MR analysis (MDD = 57) (referred to as restricted MVMR onwards; described in the supplement). There were 49 BMI SNPs after pruning and a final exposure instrument of 99 SNPs for the Depression-GlycA MVMR. After harmonization with GlycA, there were 47 SNPs available for the MVMR analysis.

##### Software

Multivariable linear regression analyses were performed using STATA version 17.0. All other analyses were performed in R Software version 4.1.1. Code for data management and statistical analysis has been made available in: daisycrick/MR_Inflammation_Depression: Investigating the bidirectional association between GlycA and MDD (github.com).

## Results

### ALSPAC cohort study

In ALSPAC, observed mean GlycA levels were 1.21 mmol/L (SD: 0.13) at age 18y and 1.22 mmol/L (SD: 0.17) at age 24y. Median SMFQ scores were 5 (Interquartile range: 3 to 9) at 18y and 5; IQR: 2 -10) at 24y. The proportions of individuals with a CIS-R depression diagnosis were 7% and 10% respectively. The distributions of observed and imputed characteristics at ages 18y and 24y are presented in supplementary table 6.

Results from the multivariable regression using the imputed data are presented in table 1. There was no evidence of crude or adjusted associations between depressive scores and GlycA. There was no evidence of any association between depression at 18y and GlycA at 24y. There was a positive relationship between GlycA levels at 18y and depression at 24y both before and after adjustment (table 2).

**Table 1:**
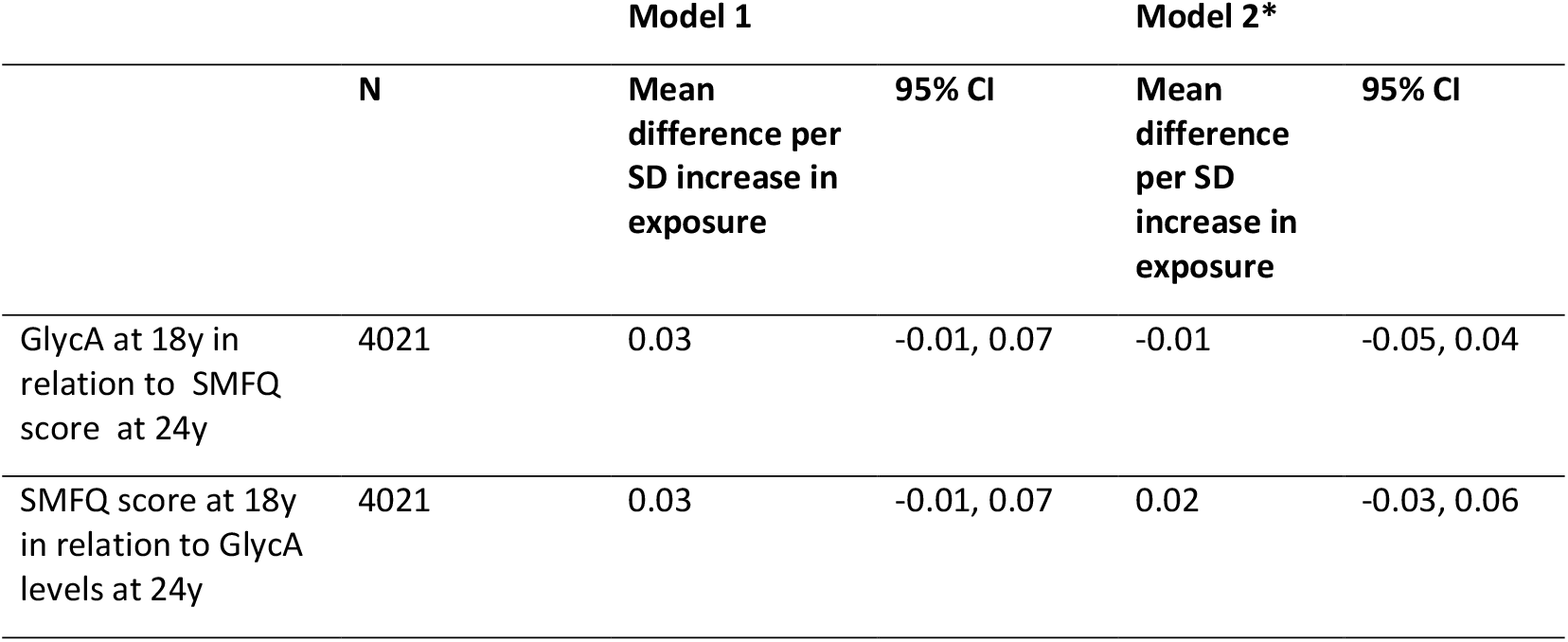
Bidirectional longitudinal associations between continuous SMFQ and GlycA. * Adjusted for smoking status, drinking status, age in months at baseline, sex, ethnicity, maternal highest education qualification and BMI at 18y.

|  | N | Model 1 |  | Model 2* |  |
| --- | --- | --- | --- | --- | --- |
|  |  | Mean difference per SD increase in exposure | 95% CI | Mean difference per SD increase in exposure | 95% CI |
| GlycA at 18y in relation to SMFQ score at 24y | 4021 | 0.03 | -0.01, 0.07 | -0.01 | -0.05, 0.04 |
| SMFQ score at 18y in relation to GlycA levels at 24y | 4021 | 0.03 | -0.01, 0.07 | 0.02 | -0.03, 0.06 |

**Table 2:** Bi-directional longitudinal association between depression and GlycA. * Adjusted for smoking status, drinking status, age in months at baseline, sex, ethnicity, maternal highest education qualification and BMI at 18y.

|  | N | Model 1 |  | Model 2 |  |
| --- | --- | --- | --- | --- | --- |
|  |  | OR | 95% CI | OR | 95% CI |
| GlycA at 18y in relation to depression at 24y | 4021 | 1.30 | 1.14, 1.47 | 1.18 | 1.03, 1.36 |
|  | N | Mean difference per SD increase in exposure | 95% CI | Mean difference per SD increase in exposure | 95% CI |
| Depression at 18y in relation to GlycA at 24y | 4021 | 0.13 | -0.03, 0.29 | 0.08 | -0.08, 0.23 |

Results from the complete case analysis are presented in supplementary table 7 and were consistent with the main findings.

### Mendelian Randomisation Analyses Instrument validity

SNP-level F statistics were all >10 in univariable analyses, indicating that IVW estimates were not subject to weak instrument bias. The total variance explained (R^2)^ across all SNPs in the univariable MR for GlycA-MDD was 4.64%, for GlycA-depressive symptoms was 3.84% and for MDD-GlycA was 0.36%. All SNPs passed Steiger filtering which shows that they explain more variation in the exposure than in the outcome for each analysis. Genetic instruments, sample sizes and instrument F-statistics for each GWAS are presented in supplementary table 8.

#### Potential Causal Effect of GlycA on genetically predicted Depression Diagnosis/ Symptoms

We found no evidence of an effect of GlycA on genetically predicted MDD risk in the MR-IVW analysis (IVW OR = 1.02, 95% CI: 0.99, 1.05). This was consistent across the other MR methods (MR-Egger, weighted median and weighted mode) (figure 1). There was no evidence of directional pleiotropic effects (MR-Egger-intercept -0.0004, p=0.805), however MR-PRESSO suggested that horizontal pleiotropy was present (p= 0.232) but identified no outlier SNPs. There was also no heterogeneity detected in either the IVW and MR-Egger analysis (Q=54.41, p=0.276 and Q=54.34, p=0.246 respectively).

**Figure 1:**
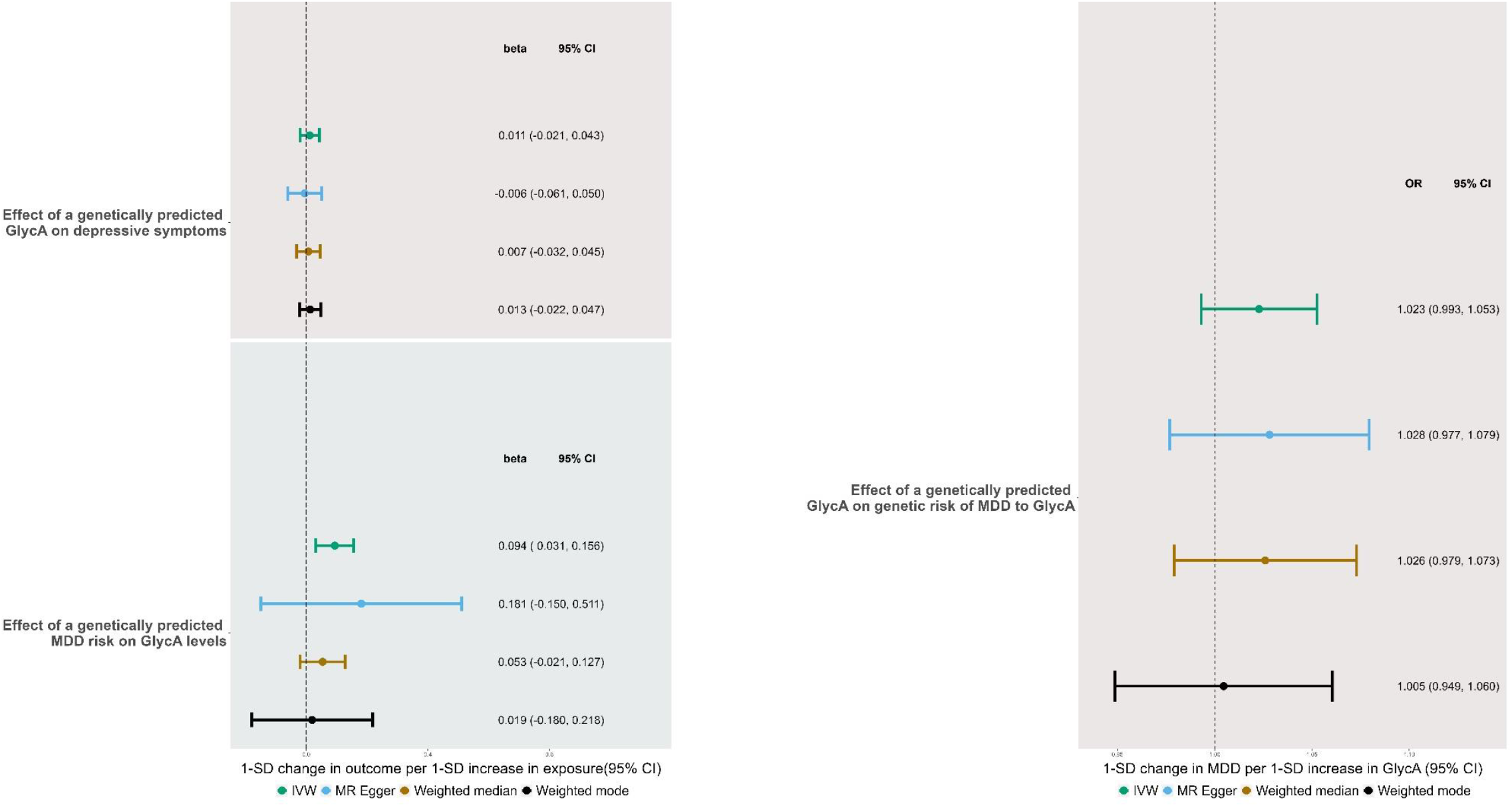
Forest Plot of IVW and sensitivity analyses showing the bidirectional relationship between genetically predicted GlycA on depressive symptoms and MDD.

We found no effect of GlycA on genetically predicted depressive symptoms (IVW mean difference in depressive symptoms per unit change in GlycA =0.01, 95% CI: -0.02, 0.04). This was consistent across the other MR methods (MR-Egger, weighted median and weighted mode) (figure 1). There was little evidence of pleiotropic effects suggested by the MR-Egger-intercept (0.001, p=0.477). MR-PRESSO found evidence for horizontal pleiotropy (p= 0.02) but identified no outlier SNPs. Heterogeneity was detected in both the IVW and MR-Egger analysis (Q=60.11, p=0.017 and Q=59.30, p=0.015 respectively). Radial MR did not change the results or identify any outliers (results in the supplement).

#### Potential Causal effect of MDD on genetically predicted GlycA Levels

We found evidence of a positive effect of MDD on genetically predicted GlycA levels in the MR-IVW analysis (mean difference in GlycA per SD increase in genetically instrumented MDD= 0.09, 95% CI = 0.03,0.16). However, other MR methods yielded estimates consistent with no effect (figure 1). There was little evidence of pleiotropic effects (MR-Egger-intercept -0.003, p =0.601) although the MR-Egger regression is imprecisely estimated. MR PRESSO suggested the presence of horizontal pleiotropy (p<0.001) and detected 1 outlier, but its removal did not alter results (comparison between original and outlier corrected results p=0.721). There was also evidence of heterogeneity in the IVW and MR-Egger analysis (Q=46.0 and 45.0 respectively, both p<0.001). Radial MR removed 1 SNP but results did not change (see supplement).

### Further Analyses

We found no effect of GlycA on depressive symptoms in MR Lap, consistent with the primary analysis. This suggests that no bias was incurred from sample overlap, winner’s curse or weak instrument bias (see supplement).

We found no effect of genetically determined IL-6 levels on MDD using the Wald ratio. We also found no effect of a genetically predisposition to MDD on IL-6 levels consistent with previous reports.^(3,7,63-66)^ See supplement and supplementary figure 2. Given that these results replicate previous findings in the literature it provides assurance of the quality in our methodology and data sources.

### MVMR controlling for the effect of BMI

For the independent effect of MDD and BMI on GlycA, the conditional F-statistic ^(70)^ suggested the instruments were strong for BMI (F=35.50) but weak for MDD (F=4.88). Genetically predicted MDD risk did not have an effect on GlycA (IVW mean difference in GlycA per SD increase in genetically instrumented MDD = 0.04, 95% CI: -0.03, 0.11) (figure 2). However, there was evidence of heterogeneity across the combined MDD and BMI instruments (MVMR Q statistic: p=5.32×10^−19^) suggesting that there could be pleiotropy in the combined set of SNPS. there was a direct effect of BMI on GlycA (IVW mean difference in GlycA per one unit change in BMI = 0.29, 95% CI: 0.25, 0.32).

**Figure 2:**
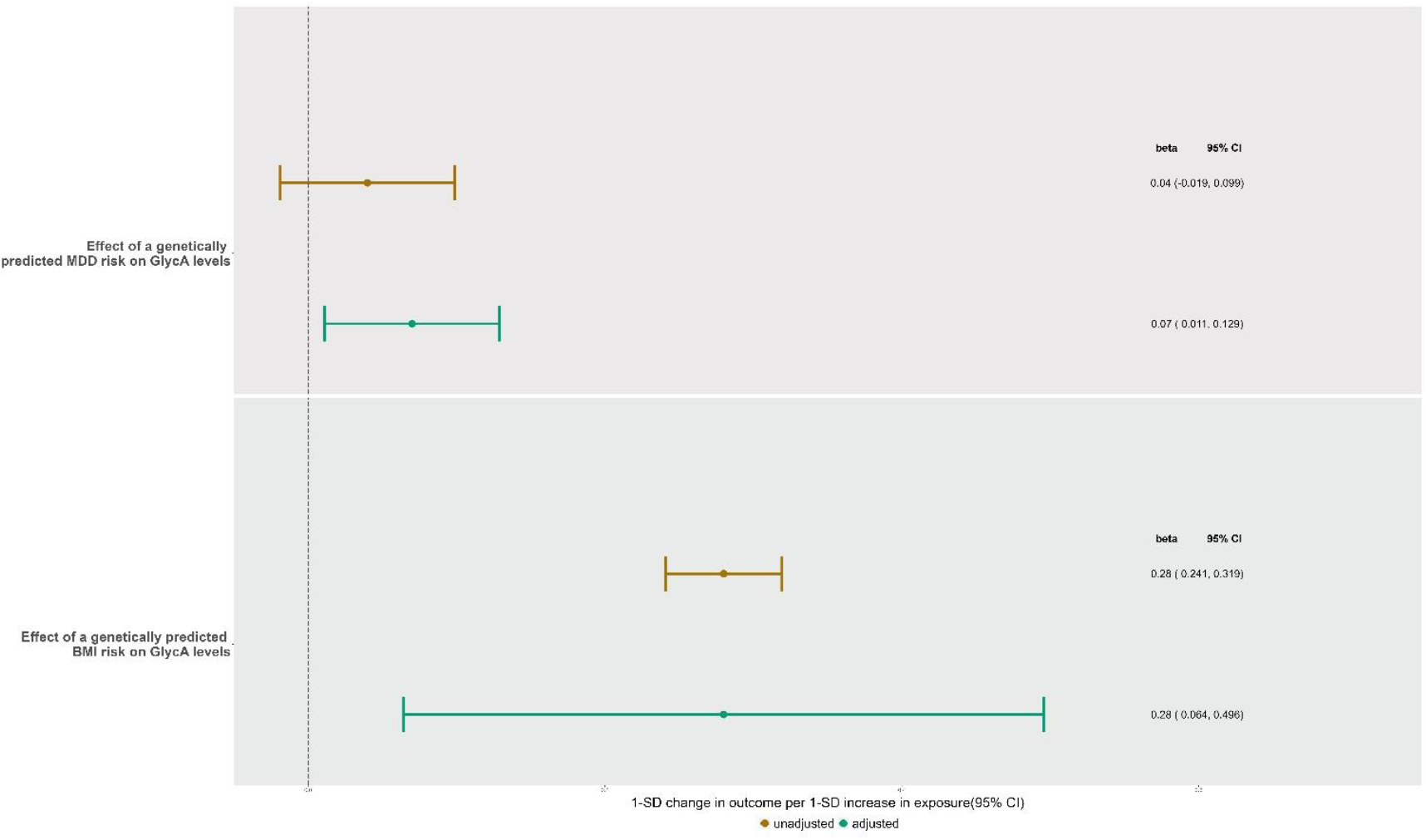
Forest Plot of MVMR-IVW estimates showing the direct effect of genetically predicted GlycA, BMI and MDD on exposures in both the unadjusted and adjusted model.

In the restricted MVMR analysis, the conditional F-statistic suggested the instruments were strong for instrument strength was good for MDD (F=28.75) but weaker for BMI (F=8.69).

There was evidence of a direct effect of MDD on GlycA (IVW = 0.07; 0.01, 0.13) (figure 2). However, again there was evidence of heterogeneity across the set of instruments (MVMR Q statistic: p=0.001). There was a direct effect of BMI on GlycA (IVW= 0.28; 0.06, 0.51).

## Discussion

We found no effects of GlycA levels on depressive symptoms or diagnosis when using multivariable regression or MR. Whilst we did find evidence for an effect of MDD on genetically predicted higher GlycA levels using MR, this did not persist in all sensitivity analyses and we did not observe an association in multivariable regression. Furthermore, when we controlled for genetically predicted BMI, through the use of MVMR, we found that the effect of MDD on genetically predicted GlycA levels attenuated to the null. Weak instrument bias may have driven this attenuation and in the restricted MVMR analysis which corrects for weak instruments, a positive effect of MDD on resultant GlycA levels persisted.

### Comparison of results to existing literature

Studies examining the links between inflammation and depression including meta-analyses of case/control studies (e.g.^(5)^), RCTs (e.g.^(71)^), longitudinal studies (e.g. ^(6)^ and cross-sectional studies (e.g. ^(72)^) have reported associations between inflammatory markers (e.g. CRP, IL-6 and TNF-alpha ^(8,21,73)^) and depression. Khandaker *et al*., found that higher levels of IL-6 in childhood were associated with risk of depression and risk of psychotic episodes in later life in a longitudinal study.^(6)^ We may have not replicated this result because inflammation exposure during sensitive periods (e.g., childhood) could play an important role in depression onset.^(74)^ As the earliest measure of GlycA we used was in adolescence, we may have missed this effect. Additional reasons we did not replicate previous findings in our multivariable regression analyses could include the different inflammatory markers studied or inflammatory marker assessment e.g. NMR signal versus immunoassay.

The majority of MR studies have focused on the effect inflammatory levels have on a genetic predisposition to MDD (e.g. ^(3,22)^). Only few have investigated the reverse association; Perry et al. ^(65)^ found no effect of a genetic predisposition to MDD on levels of immunological proteins such as IL-6. Here, we show that there may be an effect of genetically predicted MDD risk on GlycA levels. Our observed null effect of inflammation on MDD could reflect how the GlycA NMR signal is a composite of acute phase proteins.^(34,75)^ As such, the different proteins may differentially effect MDD giving an overall null result (e.g. CRP has been found to have protective effects on depression).^(21)^

## Strengths and Limitations

Here we investigated the understudied bidirectional association between inflammation and MDD using a measure of chronic systemic inflammation. ^(76)^ We used two different epidemiological methods to investigate the relationship of interest: 1) a multivariable regression longitudinal analysis using a large-population-based cohort and 2) an MR analysis, both of which can reduce bias from reverse causation. We undertook extensive sensitivity analyses and explored pleiotropic paths via BMI in our MR analyses. Future research should explore this relationship as evidence of an MDD-inflammation effect could have clinical relevance: MDD treatments may need to include anti-inflammatory aspects to combat co-morbid inflammatory health conditions.

However, ALSPAC suffers from attrition which can introduce selection bias and potentially attenuate the results towards the null. We used multiple imputation to minimise any such bias. The GWAS used were only conducted in individuals of European decent which limits the generalisability to other populations. Despite this, we had substantial power due to using summary data from large GWAS consortia. There was sample overlap between the GlycA and MDD GWAS, but the bias incurred through sample overlap is less substantial compared to biases produced by weak instruments or winner’s curse. ^(62)^ Thus we used the largest MDD GWAS to date and aimed to combat the bias using MR-lap, which found no evidence of biased effects.

## Conclusion

In conclusion, we found that inflammation as measured by GlycA did not appear to be associated with depressive diagnosis/symptoms score in both multivariable regression and MR analyses. We did find a potential causal effect of MDD on GlycA levels, but this attenuated to the null when accounting for BMI in the MVMR. However, when using the restricted MVMR which accounts for weak instrument bias, the effect returned. In light of inconsistent evidence regarding the potential associations between inflammation and depression, further work is needed to determine if reported associations are causal or not and indeed whether depression increases inflammation, vice versa, both or perhaps neither.

## Data Availability

All genetic data are available online at https://gwas.mrcieu.ac.uk/
The process of accessing ALSPAC data and samples in detail are provided on the ALSPAC website: https://www.bristol.ac.uk/alspac/researchers/access/

## Acknowledgements

This work was carried out using the computational facilities of the Advanced Computing Research Centre, University of Bristol - http://www.bristol.ac.uk/acrc/.

We are extremely grateful to all the families who took part in this study, the midwives for their help in recruiting them, and the whole ALSPAC team, which includes interviewers, computer and laboratory technicians, clerical workers, research scientists, volunteers, managers, receptionists and nurses.

## Funding

The UK Medical Research Council and Wellcome (Grant ref: 217065/Z/19/Z) and the University of Bristol provide core support for ALSPAC. This publication is the work of the authors and DC will serve as guarantor for the contents of this paper. A comprehensive list of grants funding is available on the ALSPAC website (http://www.bristol.ac.uk/alspac/external/documents/grant-acknowledgements.pdf); This research was specifically funded by Wellcome Trust and MRC (core) (Grant ref: 76467/Z/05/Z), MRC (Grant ref: MR/L022206/1) and Welllcome Trust (Grant ref: 8426812/Z/07/Z).

This work was supported in part by the GW4 BIOMED DTP (D.C., MR/N0137941/), awarded to the Universities of Bath, Bristol, Cardiff and Exeter from the Medical Research Council (MRC)/UKRI.

GMK acknowledges funding support from the Wellcome Trust (grant code: 201486/Z/16/Z), the MQ: Transforming Mental Health (grant code: MQDS17/40), the Medical Research Council UK (grant code: MC_PC_17213 and MR/S037675/1) and the BMA Foundation (J Moulton grant 2019).

AF, GK and others works in the MRC Integrative Epidemiology Unit (MC_UU_00011)

ARC and MCB work in the MRC Integrative Epidemiology Unit (MC_UU_00011/6) and are supported by the University of Bristol British Heart Foundation Accelerator Award (AA/18/7/34219)

The research by MCB was supported by a UK Medical Research Council (MRC) Skills Development Fellowship (MR/P014054/1) and a Vice-Chancellor’s Fellowship from the University of Bristol.

