## Supplementary file for "Glycoprotein Acetyls and Depression: testing for directionality and potential causality using longitudinal data and Mendelian randomization analyses"

***Contents page***

| ***Alspac Cohort recruitment*** | ***2*** |
| --- | --- |
| ***ALSPAC measures*** | ***3*** |
| ***Mendelian Randomization*** | ***5*** |
| ***Data Sources and SNP selection for main analysis*** | ***6*** |
| ***Plausibility of MR assumptions*** | ***8*** |
| ***MR-Lap*** | ***9*** |
| ***MVMR*** | ***10*** |
| ***Radial MR*** | ***12*** |
| ***IL-6 Analysis*** | ***13*** |

***Alspac Cohort Recruitment***

Please note that the ALSPAC website contains details of all the data that is available through a fully searchable data dictionary and variable search tool http://www.bristol.ac.uk/alspac/researchers/our-data/.

Ethical approval for the study was obtained from the ALSPAC Ethics and Law Committee and the Local Research Ethics Committee and can be found here: <http://www.bristol.ac.uk/alspac/researchers/research-ethics/>. Informed consent for the use of data collected via questionnaires and clinics was obtained from participants following the recommendations of the ALSPAC Ethics and Law Committee at the time. Consent for biological samples has been collected in accordance with the Human Tissue Act (2004).

Study data were collected and managed using REDCap electronic data capture tools hosted at the University of Bristol. REDCap (Research Electronic Data Capture) is a secure, web-based software platform designed to support data capture for research studies.^(1)^

**ALSPAC Measures**

**CIS-R**

The CIS-R is a diagnostic tool for depression according to the International Statistical Classification of Diseases and Related Health Problems (ICD-10). ^(2)^ It categorises individuals into 4 groups: no depression, mild depression, moderate depression and severe depression. To create our binary variable, we dichotomized these groups into: Yes/No depression diagnosis.

**GlycA**

Participants fasted overnight (or >6 hours if being seen in the afternoon) before attending the clinic. Samples were stored at -80 °C and slowly thawed in a refrigerator (+4^0^) the night before processing. The samples were mixed and spun in a centrifuge at 3400 x *g* to remove precipitation. There was no other freeze-thaw cycles and samples were analysed within 3–9 months of collection.

**Choice of primary measures**

We chose to use the SMFQ as the primary measure of depressive symptoms because as stated in the article it is a widely validated tool to measure depressive symptoms in clinical and non-clinical participants ^(3-6)^. Additionally, the SMFQ has performed well in cases and non-cases of MDD at age 25y, supporting its use in our dataset. ^(3)^ The English version has been translated into Arabic, Spanish and Norwegian and given it is a continuous measure it has greater power than the CIS-R diagnostic tool. Additionally, the SMFQ is freely available and takes approximately three-five minutes to complete ^(7)^. The SMFQ is based on a scoring system where increase in score suggests higher depressive symptoms. There is a recognised clinical cut-off of ≥12 which equates to depression ^(8)^.

GlycA was chosen as the primary measure of inflammation because we are interested in the association between chronic, systemic inflammation and depression. Given the high rates of stability seen in GlycA ^(9)^ we opted for this biomarker over other previously used inflammatory biomarkers such as C-reactive Protein or Interleukin-6. GlycA is one of the 200 metabolic measures that is quantified by NMR spectroscopy. This quantitative metabolomics platform has widespread use ^(10)^ and the all-in-one approach means that it is relatively cost-effective. Further, datasets with metabolomic data are likely to have measures of GlycA. Given the novelty of GlycA the clinical meaning of a change in GlycA score is uncertain, however higher levels of GlycA indicate higher levels of systemic chronic inflammation.

**Confounders**

To be classified as an infrequent smoker/drinker, participants had to report smoking/drinking less than once a week. To be classified as a frequent smoker/drinker, participants has to report smoking/drinking more than once a week. This variable was derived from BMI was calculated as weight in kilograms divided by height in meters squared, where weight was measured with the use of Tanita scales to the nearest 0.1 kg and height was measured using a Harpenden standiometer to the nearest 0.1 cm.

**Mendelian Randomization**

Mendelian Randomization is bound by three assumptions: (i) the genetic variants are statistically strongly associated with the exposure of interest and relevant to the population to which inference is being made (the relevance assumption); (ii) there is no confounding of the SNP-outcome association (the independence assumption); and (iii) any effect of the genetic instrument on the outcome is only via the exposure (the exclusion restriction assumption) [15] (supplementary figure 3).

***Data sources and SNP Selection for GlycA***

We obtained GlycA summary genetic estimates from a GWAS using UKBB. This was made up of 115,078 males and females from European descent. A total of 61 independent Single Nucleotide Polymorphisms (SNPs) reached genome-wide significance (p<5x10^-8^) and were selected as instrumental variables (IVs) for GlycA. Among the 61 GlycA-associated SNPs, 4 were removed due to linkage disequilibrium (r2=0.01) leaving 57 SNPs available for harmonisation.

Among the 57 GlycA-associated SNPS, 46 were available in the depression GWAS and 5 of the missing SNPs were replaced by suitable proxies. Of these remaining SNPs, 1 was excluded for being palindromic with intermediate allele frequencies. This gave 50 SNPs as instrumental variables in the analysis of GlycA on the genetic predisposition to MDD.

When harmonizing the 57 GlycA-associated SNPs with depressive symptoms, 40 SNPs were available in the dataset of depressive symptoms and 2 of the missing SNPs were replaced by suitable proxies. Of these remaining SNPs, 2 were excluded for being palindromic with intermediate allele frequencies. This resulted in 40 SNPs as instrumental variables in the analysis of GlycA to depressive symptoms.

***Data sources and SNP Selection for MDD***

This GWAS included meta-analysed data from three of the largest GWASs of depression: an MDD GWAS using 23andme data ^(11)^, an MDD GWAS using UKBB data ^(12)^ and an MDD GWAS using PGC data ^(13)^. The meta-analysed data was made up of 807,553 individuals of European-decent ^(14)^. For our analysis the GWAS from 23andme was removed due to poor MDD outcome definition (23andme use a self-reported MDD measure, compared to PGC and UKBB where MDD is based on the DSM (Diagnostic and Statistical Manual of Mental Disorders) case definition and/or validated clinical questionnaires). Therefore the final sample was made up of 500,199 males and females of European-decent and a total of 50 independent SNPs that reached genome-wide significance (p <5 x10^-8^). Of the 50 depression-associated SNPs, 1 was removed due to linkage disequilibrium (r^2^=0.01) leaving 49 SNPs available for harmonisation.

Among the 49 MDD-associated SNPs, 2 SNPs were excluded for being palindromic with intermediate allele frequencies, resulting in 47 SNPs as IVs in the analysis of MDD on GlycA. No SNPs were removed for the analysis of MDD on GlycA , giving 49 SNPs to be used as IVs.

***Data sources and SNP Selection for depressive symptoms***

We obtained genetic summary estimates for individuals with depressive symptoms from a GWAS using SSGAC. This was made up of 161,460 individuals of European-decent ^(15)^. Only 2 SNPs (p <5 x10^-8^; Linkage disequilibrium r^2^=0.01) were selected as instrumental variables for depressive symptoms. In the MR investigating a genetic liability of depressive symptoms on GlycA no proxy SNPs were required. However, 1 SNP was removed due to being palindromic with intermediate allele frequencies leaving only 1 SNP available for the analysis between depressive symptoms and GlycA. An MR was therefore not run on this association.

**Plausibility of MR assumptions**

The IVW assumes no unbalanced horizontal pleiotropy and forces the intercept to go through zero. In contrast, MR-Egger does not force the line through zero and as a result the intercept gives an indication of the presence of pleiotropy ^(16)^[45]. The weighted median and weighted mode assume that the pleiotropic effect of certain SNPs on the outcome are less likely to converge on a common median or modal estimate. However, the valid SNPs that display no pleiotropic effects will show more uniform and homogenous effects on the exposure and outcome. This makes them more likely to cluster toward the median/modal point estimate ^(17,18)^.

***MR-LAP***

MR-Lap is a novel method corrects for sample overlap, weak instrument bias and winner’s curse ^(19)^. If the MR-LAP corrected effect does not significantly differ from the observed effect, then the IVW-MR estimate can be used. If there is a difference between the two estimates, it suggests that the biases are having an effect on the effect estimate and therefore, the corrected effect would be preferred ^(19)^. For this analysis, we used the same GlycA and depressive symptoms GWAS as used in the main analysis. Instrumental variables (IVs) were pruned to the distance threshold of 10,000 Kb with an LD threshold of 0.001, giving 49 IVs as part of the analysis.

**MR-Lap results**

There was no effect of GlycA on depressive symptoms (mean difference in depression symptoms per one-unit increase of GlycA was 0.02, 95% CI: -0.01, 0.04). The estimate corrected for Winner’s curse, sample overlap and weak instrument bias was similar (mean difference in depressive symptoms per one unit increase of GlycA being 0.02; 95% CI: -0.02, 0.05). This suggests that these biases were not affecting our results (p=0.841) ^(19)^.

**Multivariable Mendelian Randomization**

Multivariable Mendelian Randomization (MVMR) estimates the direct effect of each exposure on the outcome and can be used to account for pleiotropic pathways. In this way the univariable MR can be thought of as the *total* *effect* in an epidemiological mediation model, whereas the MVMR is analogous to the *direct* *effects* of each exposure on the outcome (supplementary figure 4). This analysis adjusts for pleiotropic effects of the exposure SNPs which act via BMI.

Genetic correlations of GlycA and BMI, and depression and BMI (r=0.334 and r=0.086 respectively) were estimated through linkage disequilibrium score regression. This was to check that the variables were genetically correlated and therefore an MVMR was applicable. As described in the main manuscript, we repeated the MVMR using only the MDD SNPs identified in the univariable MR (known as the adjusted MVMR). This was because in the initial MVMR analysis, after the MDD and BMI SNPs were combined, there was a limited number of remaining MDD SNPs. This means that many of the MDD SNPs were in LD with BMI, suggesting that they may be weak instruments or that many of the MDD SNPs were acting through BMI. We did this by merging the 57 MDD SNPs with the IVs for BMI, LD clumped the combined data and then harmonised with the GlycA data.

In the MVMR analysis of MDD on GlycA, while controlling for BMI, 49 of 365 SNPs were removed due to LD or absence from LD reference panel and 32 of the missing SNPs were replaced by suitable proxies. No proxies were needed for the outcome data. During harmonization, 10 SNPs were dropped for being palindromic with intermediate allele frequencies. In the MVMR analysis of GlycA on MDD, while controlling for BMI, 69 of the 376 SNPs were removed due to LD with other SNPS or absence from LD reference panel and 2 of the missing BMI SNPs were replaced by suitable proxies. In the GlycA data, 33 of the missing SNPs were replaced by suitable proxies. During harmonization, 10 SNPs were dropped for being palindromic with intermediate allele frequencies.

Heterogeneity of instrument effects was evaluated using a modified form of Cochran’s Q statistic. This assesses horizontal pleiotropy with respect to differences in MVMR estimates across the set of instruments. Any observed heterogeneity is indicative of a violation of the exclusion restriction assumption.

Instrument strength was evaluated through a conditional F-statistic (the same conventional instrument strength threshold of 10 can be used) ^(20)^. The F-statistic assessing instrument strength, whether the genetic variants used as instruments are required to strongly associate with their exposures, conditioning on the remaining included exposures. We required the variance-covariance for the effects of the genetic variants on each exposure to test for weak instrument and heterogeneity and therefore assessed the phenotypic correlation between exposures (GlycA-BMI overlap = -0.13, Depression-BMI overlap = 0.05).

**Radial MR**

Radial MR is similar to IVW, but detects and removes outlying variants via a simulation-based approach to then re-estimate the original exposure-outcome relationship. This can reduce bias in MR estimates, but can sometimes lead to over-fitting because the standard error is reduced. Radial plots can help to interpret the validity of the IVW and MR-Egger regression and improve the detection of outliers and data points that are influential in the IVW or MR-Egger analysis.

***Effect of genetically determined GlycA levels on predisposition to depressive symptoms***

No significant outliers were detected and we observed a consistent effect direction as the original MR analysis for Radial IVW (*beta=0.01*; 95% CI -0.02, 0.04; *P* value = 0.491) and Radial MR-Egger (*beta=* -0.002; 95% CI -0.06, 0.06); *P* value =  0.959). Cochran’s Q statistic and the Rucker’s Q statistic indicated heterogeneity (p = 0.017 and p = 0.018 respectively). The radial IVW plot is presented in figure 5.

**Effect of genetic predisposition to MDD on genetically determined GlycA levels**

Using Radial-IVW, one outlier was detected and removed (rs7725715, p = 0.001). The corrected effect was consistent with the main analysis (*beta=0.09*; 95% CI 0.03, 0.15; *P* value = 0.002). The same outlier was detected (p=0.001) in the Radial MR-Egger n (*beta=* 0.23; 95% CI -0.25, 0.71]; *P* value =  0.346). It is worth noting that the Radial MR-Egger approach often has wider confidence intervals due to it being statistically relatively inefficient. Cochran’s Q statistic and the Rucker’s Q statistic indicated heterogeneity (p <0.001 for both). The radial IVW plot is presented in figure 6.

**IL-6 Analysis**

***Data sources and SNP Selection for IL-6***

We obtained IL-6 summary genetic estimates from a 26 cohort GWAS meta-analysis ^(21)^. This was the most recent and one of the largest IL-6 GWAS’ to date and made up of 52,654 males and females from European descent. They identified 94 variants that were genome-wide significantly associated with IL-6 levels. However, only 2 SNPs (p <5 x10^-8^; Linkage disequilibrium r^2^=0.01) were selected as instrumental variables due to high levels of LD. In the MR investigating a genetic liability of IL-6 on MDD one proxy SNP were not available leaving only 1 SNP available for the analysis. Previous analyses using IL-6 exposure IVs has reported a similar issue and we therefore opted to use a Wald ratio for this analysis which will complement previous work ^(22)^.

We used the same MDD GWAS as in the main MR analysis with GlycA and therefore had 49 MDD-associated SNPs. Among the 49 MDD-associated SNPs, 1 SNP was excluded for being palindromic with intermediate allele frequencies. No proxies were available, resulting in 22 SNPs as IVs in the analysis of MDD on IL-6. Harmonized SNPs are presented in Supplementary tables 9-10.

***Potential Causal Effect of IL-6 on Depression Diagnosis***

We found no effect of IL-6 on genetically predicted risk of MDD (Wald Ratio OR = 1.10, 95% CI: 0.94, 1.28) (supplementary figure 2). The F statistic suggested that the instrument was weak and Steiger filtering suggested that reverse causation was likely, but due to being the only remaining SNP we did not remove it from the analysis.

***Potential Causal effect of depression diagnosis on IL-6 Levels***

We found no effect MDD risk on IL-6 levels in the MR-IVW analysis (mean difference in IL-6 per SD increase in genetically instrumented MDD= 0.01, 95% CI = -0.09,0.01). This was consistent across the other MR methods (MR-egger, weighted median and weighted mode) (figure 2). Under the assumption that pleiotropy is independent of the SNP-exposure association there was little evidence of pleiotropic effects suggested by the MR-Egger-intercept (-0.0001, p=0.988). MR-PRESSO did not detect any outliers and the MR PRESSO Global test found no suggestion of horizontal pleiotropy for the effect of IL-6 on MDD (p=0.677) and heterogeneity was not detected in either the IVW and MR-Egger analysis (Q=10, p=1.00 and Q=9, p=1.00 respectively). Of the 22 SNPs, 11 did not pass steiger filtering. However, the results did not change and we present results with the SNPs removed.

19.	Mounier N, Kutalik ZJb. Correction for sample overlap, winner’s curse and weak instrument bias in two-sample Mendelian Randomization. 2021.

20.	Sanderson E, Spiller W, Bowden J. Testing and correcting for weak and pleiotropic instruments in two‐sample multivariable Mendelian randomization. Statistics in Medicine. 2021;40(25):5434-52.

21.	Ahluwalia TS, Prins BP, Abdollahi M, Armstrong NJ, Aslibekyan S, Bain L, et al. Genome-wide association study of circulating interleukin 6 levels identifies novel loci. Hum Mol Genet. 2021;30(5):393-409.

22.	Perry BI, Upthegrove R, Kappelmann N, Jones PB, Burgess S, Khandaker GM. Associations of immunological proteins/traits with schizophrenia, major depression and bipolar disorder: A bi-directional two-sample mendelian randomization study. Brain Behav Immun. 2021;97:176-85.
