## Supplementary tables for "Glycoprotein Acetyls and Depression: testing for directionality and potential causality using longitudinal data and Mendelian randomization analyses"

| **Main Variables** |
| --- |
| GlycA levels at 18y |
| GlycA levels at 24y |
| SMFQ score at 18y |
| SMFQ score at 24y |
| CIS-R diagnosis at age 18y |
| CIS-R diagnosis at age 24y |
| Age in months at 18y clinic |
| Age in months at 24y clinic |
| BMI at 18y clinic |
| BMI at 24y clinic |
| Ethnicity |
| Sex |
| Type of drinker at 24y clinic |
| Type of drinker at 18y clinic |
| Type of smoker at 24y clinic |
| Type of smoker at 18y clinic |
| Maternal highest education qualification |
| **Auxiliary Variables** |
| GlycA levels at age 7y |
| GlycA levels at age 15y |
| Moods and Feelings questionnaire at t8 |
| Moods and Feelings questionnaire at t7 |
| Moods and Feelings questionnaire at t6 |
| Moods and Feelings questionnaire at t4 |
| Moods and Feelings questionnaire at t3 |
| Moods and Feelings questionnaire at t9 |
| C-Reactive Protein levels at 9y clinic |
| C-Reactive Protein levels at 15y clinic |
| C-Reactive Protein levels at 24y clinic |
| Interleukin-6 levels at 9y clinic |
| Classic ACE score |
| Extended ACE score |
| Maternal social class |
| Maternal smoking status when pregnant |
| Household highest social class |
| **Supplementary Table 1: Main and auxiliary variables included to help inform the imputation** |

| **SNP** | **Effect Allele** | **Other Allele** | **beta** | **EAF** | **se** | ***p*** | **R^2^** | **F statistic** | **Steiger Direction** | **Steiger *p*** |
| --- | --- | --- | --- | --- | --- | --- | --- | --- | --- | --- |
| rs10455872 | G | A | -0.08 | 0.08 | 0.008 | 1.00E-25 | 0.001 | 104.14 | TRUE | 1.74E-20 |
| rs112875651 | A | G | -0.07 | 0.39 | 0.004 | 7.00E-65 | 0.002 | 277.81 | TRUE | 1.28E-49 |
| rs113354603 | A | G | 0.13 | 0.07 | 0.009 | 1.00E-50 | 0.002 | 245.53 | TRUE | 2.31E-40 |
| rs1168032 | G | A | 0.04 | 0.64 | 0.004 | 2.30E-20 | 0.001 | 77.20 | TRUE | 6.22E-15 |
| rs116843064 | A | G | -0.10 | 0.02 | 0.015 | 4.00E-11 | 0.000 | 43.71 | TRUE | 2.65E-08 |
| rs117155836 | A | G | 0.14 | 0.03 | 0.012 | 6.20E-30 | 0.001 | 122.58 | TRUE | 1.67E-20 |
| rs117733303 | G | A | -0.13 | 0.02 | 0.015 | 3.70E-17 | 0.001 | 68.59 | TRUE | 3.84E-14 |
| rs12032372 | C | T | -0.03 | 0.21 | 0.005 | 4.70E-09 | 0.0003 | 32.74 | TRUE | 3.11E-07 |
| rs12073837 | T | C | -0.03 | 0.28 | 0.005 | 1.60E-08 | 0.0003 | 30.43 | TRUE | 4.40E-06 |
| rs1260326 | C | T | -0.10 | 0.60 | 0.004 | 2.60E-125 | 0.005 | 559.31 | TRUE | 1.63E-99 |
| rs13108218 | G | A | -0.04 | 0.62 | 0.004 | 1.60E-19 | 0.001 | 79.25 | TRUE | 8.44E-16 |
| rs143578173 | T | C | -0.14 | 0.03 | 0.013 | 3.40E-27 | 0.001 | 123.79 | TRUE | 6.28E-21 |
| rs144018203 | C | G | 0.12 | 0.01 | 0.021 | 1.20E-08 | 0.0003 | 34.87 | TRUE | 2.92E-07 |
| rs146203232 | T | C | 0.05 | 0.07 | 0.008 | 5.30E-09 | 0.0003 | 33.97 | TRUE | 1.51E-07 |
| rs149807892 | T | C | -0.12 | 0.02 | 0.017 | 8.80E-14 | 0.0005 | 54.09 | TRUE | 7.79E-11 |
| rs150844304 | C | A | 0.07 | 0.03 | 0.013 | 4.40E-08 | 0.0003 | 29.74 | TRUE | 1.05E-06 |
| rs151083835 | G | C | 0.10 | 0.02 | 0.015 | 2.00E-12 | 0.0004 | 47.75 | TRUE | 4.60E-09 |
| rs17105232 | A | G | 0.03 | 0.25 | 0.005 | 1.40E-10 | 0.0003 | 39.59 | TRUE | 1.23E-08 |
| rs17580 | A | T | -0.05 | 0.05 | 0.010 | 8.80E-09 | 0.0002 | 27.97 | TRUE | 5.64E-05 |
| rs1801689 | C | A | -0.07 | 0.03 | 0.012 | 2.40E-10 | 0.0003 | 36.25 | TRUE | 4.09E-07 |
| rs182050989 | T | C | 0.07 | 0.03 | 0.012 | 1.50E-08 | 0.0003 | 33.54 | TRUE | 4.19E-07 |
| rs2070634 | G | T | 0.04 | 0.51 | 0.004 | 2.10E-19 | 0.001 | 81.11 | TRUE | 9.60E-14 |
| rs2294915 | T | C | -0.03 | 0.23 | 0.005 | 3.60E-11 | 0.0004 | 43.74 | TRUE | 2.34E-09 |
| rs2445818 | G | A | 0.05 | 0.93 | 0.008 | 2.60E-08 | 0.0003 | 29.48 | TRUE | 5.41E-06 |
| rs2497337 | T | C | 0.02 | 0.51 | 0.004 | 1.40E-08 | 0.0003 | 30.22 | TRUE | 1.40E-06 |
| rs28929474 | T | C | -0.28 | 0.02 | 0.015 | 3.80E-80 | 0.003 | 352.77 | TRUE | 1.62E-58 |
| rs325 | C | T | -0.09 | 0.10 | 0.007 | 5.60E-36 | 0.001 | 151.80 | TRUE | 1.93E-26 |
| rs4297946 | C | G | 0.02 | 0.47 | 0.004 | 1.50E-08 | 0.0003 | 30.26 | TRUE | 2.64E-04 |
| rs4459081 | C | G | 0.03 | 0.65 | 0.004 | 3.70E-15 | 0.001 | 58.51 | TRUE | 4.38E-10 |
| rs55780214 | A | T | -0.03 | 0.29 | 0.005 | 4.40E-12 | 0.0004 | 45.10 | TRUE | 3.05E-09 |
| rs56188865 | C | T | -0.03 | 0.37 | 0.004 | 3.10E-11 | 0.0004 | 43.58 | TRUE | 4.57E-09 |
| rs58542926 | T | C | -0.06 | 0.07 | 0.008 | 7.80E-13 | 0.0004 | 49.62 | TRUE | 3.78E-08 |
| rs59296513 | A | G | -0.03 | 0.32 | 0.004 | 1.80E-09 | 0.0003 | 32.64 | TRUE | 3.71E-06 |
| rs59774409 | T | C | 0.08 | 0.08 | 0.007 | 3.30E-28 | 0.001 | 121.58 | TRUE | 5.00E-22 |
| rs62128087 | T | C | -0.04 | 0.16 | 0.006 | 3.30E-13 | 0.0005 | 53.68 | TRUE | 9.71E-10 |
| rs62259939 | A | G | 0.03 | 0.43 | 0.004 | 1.10E-09 | 0.0003 | 37.05 | TRUE | 1.38E-06 |
| rs62466318 | T | C | -0.09 | 0.20 | 0.005 | 8.50E-63 | 0.002 | 276.10 | TRUE | 1.05E-48 |
| rs6424109 | A | C | 0.04 | 0.87 | 0.006 | 1.40E-08 | 0.0003 | 32.22 | TRUE | 1.42E-06 |
| rs6452937 | G | A | -0.03 | 0.14 | 0.006 | 6.20E-09 | 0.0002 | 28.45 | TRUE | 1.68E-06 |
| rs6601299 | C | T | 0.07 | 0.90 | 0.007 | 2.90E-23 | 0.001 | 97.24 | TRUE | 1.44E-17 |
| rs6717858 | C | T | -0.02 | 0.40 | 0.004 | 3.80E-08 | 0.0002 | 25.98 | TRUE | 2.69E-05 |
| rs6734238 | G | A | 0.03 | 0.40 | 0.004 | 4.00E-09 | 0.0003 | 34.64 | TRUE | 1.04E-07 |
| rs676210 | A | G | -0.03 | 0.21 | 0.005 | 2.20E-08 | 0.0003 | 30.92 | TRUE | 3.07E-06 |
| rs687621 | G | A | 0.03 | 0.32 | 0.004 | 6.30E-11 | 0.0004 | 40.95 | TRUE | 2.00E-07 |
| rs72801474 | A | G | -0.04 | 0.09 | 0.007 | 1.10E-08 | 0.0003 | 32.91 | TRUE | 6.26E-07 |
| rs7697204 | T | C | 0.03 | 0.74 | 0.005 | 3.20E-11 | 0.0003 | 40.20 | TRUE | 3.08E-07 |
| rs77303550 | T | C | 0.16 | 0.19 | 0.005 | 1.00E-200 | 0.008 | 968.85 | TRUE | 3.90E-168 |
| rs78689694 | C | G | 0.03 | 0.13 | 0.006 | 4.70E-08 | 0.0003 | 31.18 | TRUE | 1.13E-06 |
| rs7924036 | T | G | -0.03 | 0.50 | 0.004 | 2.80E-12 | 0.0004 | 43.73 | TRUE | 2.12E-08 |
| rs79287178 | A | G | 0.10 | 0.03 | 0.012 | 7.70E-16 | 0.001 | 69.10 | TRUE | 4.10E-11 |
| rs964184 | C | G | -0.11 | 0.87 | 0.006 | 2.70E-68 | 0.003 | 295.06 | TRUE | 9.42E-53 |
| **Supplementary Table 2: Harmonised SNPs for the association between GlycA and MDD with exposure information** | | | | | | | | | | |

| **SNP** | **Effect Allele** | **Other Allele** | **beta** | **eaf** | **se** | ***p*** | **F** | **R^2^** | **steiger direction** | **steiger *p*** |
| --- | --- | --- | --- | --- | --- | --- | --- | --- | --- | --- |
| rs1021363 | G | A | -0.03 | 0.64 | 0.005 | 2.29E-11 | 206.66 | 8.88E-05 | TRUE | 0.84 |
| rs10235664 | C | T | -0.03 | 0.25 | 0.005 | 4.68E-08 | 137.83 | 6.07E-05 | TRUE | 0.52 |
| rs10913112 | T | C | -0.03 | 0.38 | 0.005 | 4.53E-09 | 161.51 | 6.78E-05 | TRUE | 0.24 |
| rs12919291 | C | G | 0.03 | 0.19 | 0.006 | 3.09E-09 | 163.62 | 7.07E-05 | TRUE | 0.39 |
| rs12967143 | C | G | -0.03 | 0.70 | 0.005 | 2.53E-13 | 249.60 | 0.000108 | TRUE | 0.00 |
| rs13037326 | T | C | 0.03 | 0.26 | 0.005 | 2.40E-10 | 184.90 | 8.00E-05 | TRUE | 0.06 |
| rs1367635 | C | T | 0.03 | 0.51 | 0.004 | 4.35E-09 | 160.00 | 6.92E-05 | TRUE | 0.02 |
| rs150186873 | C | A | 0.07 | 0.03 | 0.012 | 4.51E-09 | 156.88 | 6.88E-05 | TRUE | 0.01 |
| rs150346963 | T | C | 0.03 | 0.41 | 0.004 | 1.16E-10 | 194.14 | 8.27E-05 | TRUE | 0.01 |
| rs17641524 | T | C | -0.03 | 0.21 | 0.005 | 1.50E-08 | 149.47 | 6.41E-05 | TRUE | 0.03 |
| rs1931388 | G | A | -0.03 | 0.40 | 0.004 | 1.68E-11 | 209.75 | 8.99E-05 | TRUE | 0.33 |
| rs1950829 | G | A | -0.03 | 0.52 | 0.004 | 4.74E-12 | 220.44 | 9.54E-05 | TRUE | 0.00 |
| rs198457 | T | C | -0.03 | 0.19 | 0.006 | 1.90E-08 | 151.95 | 6.33E-05 | TRUE | 0.35 |
| rs2111592 | A | G | 0.03 | 0.31 | 0.005 | 1.35E-08 | 149.12 | 6.53E-05 | TRUE | 0.03 |
| rs2214123 | G | A | -0.03 | 0.65 | 0.005 | 8.56E-09 | 155.77 | 6.72E-05 | TRUE | 0.06 |
| rs2232423 | G | A | -0.06 | 0.11 | 0.007 | 1.14E-18 | 363.47 | 0.000157 | TRUE | 0.35 |
| rs2418449 | C | T | -0.03 | 0.28 | 0.005 | 4.25E-09 | 159.65 | 6.85E-05 | TRUE | 0.38 |
| rs247910 | G | A | 0.02 | 0.46 | 0.004 | 4.71E-08 | 139.48 | 6.07E-05 | TRUE | 0.14 |
| rs2522831 | C | T | 0.02 | 0.47 | 0.004 | 2.11E-08 | 143.71 | 6.23E-05 | TRUE | 0.03 |
| rs2568958 | A | G | 0.04 | 0.60 | 0.004 | 2.90E-18 | 349.35 | 0.000151 | TRUE | 0.00 |
| rs28541419 | G | C | -0.03 | 0.23 | 0.005 | 1.76E-08 | 151.48 | 6.30E-05 | TRUE | 0.31 |
| rs2876520 | G | C | 0.03 | 0.47 | 0.004 | 2.24E-09 | 168.47 | 7.31E-05 | TRUE | 0.01 |
| rs30266 | A | G | 0.04 | 0.33 | 0.005 | 1.43E-15 | 295.13 | 0.000127 | TRUE | 0.04 |
| rs354155 | C | G | -0.04 | 0.09 | 0.008 | 1.75E-09 | 169.03 | 7.16E-05 | TRUE | 0.06 |
| rs3807865 | A | G | 0.03 | 0.41 | 0.004 | 1.09E-12 | 232.75 | 9.92E-05 | TRUE | 0.08 |
| rs4141983 | C | T | -0.03 | 0.33 | 0.005 | 9.69E-09 | 153.25 | 6.58E-05 | TRUE | 0.03 |
| rs4497414 | C | T | 0.03 | 0.44 | 0.004 | 2.93E-11 | 208.82 | 8.74E-05 | TRUE | 0.12 |
| rs4730387 | A | T | 0.02 | 0.47 | 0.004 | 4.12E-08 | 141.05 | 6.12E-05 | TRUE | 0.18 |
| rs4799949 | T | C | -0.03 | 0.67 | 0.005 | 1.40E-10 | 189.13 | 8.06E-05 | TRUE | 0.38 |
| rs4936276 | C | G | 0.03 | 0.62 | 0.004 | 3.57E-10 | 181.84 | 7.98E-05 | TRUE | 0.47 |
| rs508502 | T | C | -0.03 | 0.30 | 0.005 | 3.56E-08 | 146.24 | 6.05E-05 | TRUE | 0.03 |
| rs59082935 | T | C | 0.04 | 0.13 | 0.007 | 3.07E-08 | 153.21 | 6.05E-05 | TRUE | 0.02 |
| rs59283172 | A | G | -0.04 | 0.11 | 0.007 | 2.41E-08 | 146.75 | 6.21E-05 | TRUE | 0.02 |
| rs61914045 | A | G | 0.03 | 0.20 | 0.005 | 7.96E-09 | 154.82 | 6.55E-05 | TRUE | 0.29 |
| rs62535714 | A | G | 0.03 | 0.16 | 0.006 | 4.69E-09 | 157.60 | 6.83E-05 | TRUE | 0.20 |
| rs66511648 | C | T | 0.03 | 0.28 | 0.005 | 6.03E-10 | 179.50 | 7.65E-05 | TRUE | 0.21 |
| rs7152906 | C | T | 0.03 | 0.52 | 0.004 | 1.87E-09 | 166.28 | 7.20E-05 | TRUE | 0.08 |
| rs7241572 | A | G | 0.03 | 0.20 | 0.005 | 2.43E-09 | 169.97 | 7.15E-05 | TRUE | 0.72 |
| rs72948506 | A | G | 0.03 | 0.30 | 0.005 | 1.71E-08 | 146.87 | 6.36E-05 | TRUE | 0.69 |
| rs7538938 | C | T | 0.03 | 0.56 | 0.004 | 7.29E-09 | 155.35 | 6.81E-05 | TRUE | 0.31 |
| rs754287 | A | T | -0.03 | 0.37 | 0.005 | 1.31E-10 | 194.05 | 8.25E-05 | TRUE | 0.18 |
| rs7551758 | G | T | 0.03 | 0.53 | 0.004 | 5.11E-11 | 199.51 | 8.66E-05 | TRUE | 0.02 |
| rs76954012 | A | T | 0.04 | 0.09 | 0.007 | 2.41E-08 | 143.42 | 6.20E-05 | TRUE | 0.49 |
| rs7725715 | A | G | 0.03 | 0.53 | 0.004 | 1.61E-11 | 209.43 | 9.09E-05 | TRUE | 0.61 |
| rs843812 | A | G | 0.02 | 0.41 | 0.004 | 1.41E-08 | 149.07 | 6.35E-05 | TRUE | 0.03 |
| rs9364755 | G | A | 0.03 | 0.23 | 0.005 | 3.49E-08 | 140.28 | 6.16E-05 | TRUE | 0.72 |
| rs9529218 | T | C | -0.03 | 0.20 | 0.005 | 2.23E-10 | 187.24 | 7.92E-05 | TRUE | 0.36 |
| rs9536381 | T | C | 0.03 | 0.33 | 0.005 | 2.62E-08 | 142.95 | 6.14E-05 | TRUE | 0.26 |
| rs9831648 | T | G | -0.03 | 0.77 | 0.005 | 1.59E-08 | 149.30 | 6.30E-05 | TRUE | 0.33 |
| **Supplementary Table 3: Harmonised SNPs for the association between GlycA and depressive symptoms with exposure information** | | | | | | | | | | |

| **SNP** | **Effect Allele** | **Other Allele** | **beta** | **eaf** | **se** | ***p*** | **F** | **R^2^** | **steiger direction** | **steiger *p*** |
| --- | --- | --- | --- | --- | --- | --- | --- | --- | --- | --- |
| rs1021363 | G | A | -0.03 | 0.64 | 0.005 | 2.29E-11 | 206.66 | 8.88E-05 | TRUE | 0.84 |
| rs10235664 | C | T | -0.03 | 0.25 | 0.005 | 4.68E-08 | 137.83 | 6.07E-05 | TRUE | 0.52 |
| rs10913112 | T | C | -0.03 | 0.38 | 0.005 | 4.53E-09 | 161.51 | 6.78E-05 | TRUE | 0.24 |
| rs12919291 | C | G | 0.03 | 0.19 | 0.006 | 3.09E-09 | 163.62 | 7.07E-05 | TRUE | 0.39 |
| rs12967143 | C | G | -0.03 | 0.70 | 0.005 | 2.53E-13 | 249.60 | 0.000108 | TRUE | 0.00 |
| rs13037326 | T | C | 0.03 | 0.26 | 0.005 | 2.40E-10 | 184.90 | 8.00E-05 | TRUE | 0.06 |
| rs1367635 | C | T | 0.03 | 0.51 | 0.004 | 4.35E-09 | 160.00 | 6.92E-05 | TRUE | 0.02 |
| rs150186873 | C | A | 0.07 | 0.03 | 0.012 | 4.51E-09 | 156.88 | 6.88E-05 | TRUE | 0.01 |
| rs150346963 | T | C | 0.03 | 0.41 | 0.004 | 1.16E-10 | 194.14 | 8.27E-05 | TRUE | 0.01 |
| rs17641524 | T | C | -0.03 | 0.21 | 0.005 | 1.50E-08 | 149.47 | 6.41E-05 | TRUE | 0.03 |
| rs1931388 | G | A | -0.03 | 0.40 | 0.004 | 1.68E-11 | 209.75 | 8.99E-05 | TRUE | 0.33 |
| rs1950829 | G | A | -0.03 | 0.52 | 0.004 | 4.74E-12 | 220.44 | 9.54E-05 | TRUE | 0.00 |
| rs198457 | T | C | -0.03 | 0.19 | 0.006 | 1.90E-08 | 151.95 | 6.33E-05 | TRUE | 0.35 |
| rs2111592 | A | G | 0.03 | 0.31 | 0.005 | 1.35E-08 | 149.12 | 6.53E-05 | TRUE | 0.03 |
| rs2214123 | G | A | -0.03 | 0.65 | 0.005 | 8.56E-09 | 155.77 | 6.72E-05 | TRUE | 0.06 |
| rs2232423 | G | A | -0.06 | 0.11 | 0.007 | 1.14E-18 | 363.47 | 0.000157 | TRUE | 0.35 |
| rs2418449 | C | T | -0.03 | 0.28 | 0.005 | 4.25E-09 | 159.65 | 6.85E-05 | TRUE | 0.38 |
| rs247910 | G | A | 0.02 | 0.46 | 0.004 | 4.71E-08 | 139.48 | 6.07E-05 | TRUE | 0.14 |
| rs2522831 | C | T | 0.02 | 0.47 | 0.004 | 2.11E-08 | 143.71 | 6.23E-05 | TRUE | 0.03 |
| rs2568958 | A | G | 0.04 | 0.60 | 0.004 | 2.90E-18 | 349.35 | 0.000151 | TRUE | 0.00 |
| rs28541419 | G | C | -0.03 | 0.23 | 0.005 | 1.76E-08 | 151.48 | 6.30E-05 | TRUE | 0.31 |
| rs30266 | A | G | 0.04 | 0.33 | 0.005 | 1.43E-15 | 295.13 | 0.000127 | TRUE | 0.04 |
| rs354155 | C | G | -0.04 | 0.09 | 0.008 | 1.75E-09 | 169.03 | 7.16E-05 | TRUE | 0.06 |
| rs3807865 | A | G | 0.03 | 0.41 | 0.004 | 1.09E-12 | 232.75 | 9.92E-05 | TRUE | 0.08 |
| rs4141983 | C | T | -0.03 | 0.33 | 0.005 | 9.69E-09 | 153.25 | 6.58E-05 | TRUE | 0.03 |
| rs4497414 | C | T | 0.03 | 0.44 | 0.004 | 2.93E-11 | 208.82 | 8.74E-05 | TRUE | 0.12 |
| rs4799949 | T | C | -0.03 | 0.67 | 0.005 | 1.40E-10 | 189.13 | 8.06E-05 | TRUE | 0.38 |
| rs4936276 | C | G | 0.03 | 0.62 | 0.004 | 3.57E-10 | 181.84 | 7.98E-05 | TRUE | 0.47 |
| rs508502 | T | C | -0.03 | 0.30 | 0.005 | 3.56E-08 | 146.24 | 6.05E-05 | TRUE | 0.03 |
| rs59082935 | T | C | 0.04 | 0.13 | 0.007 | 3.07E-08 | 153.21 | 6.05E-05 | TRUE | 0.02 |
| rs59283172 | A | G | -0.04 | 0.11 | 0.007 | 2.41E-08 | 146.75 | 6.21E-05 | TRUE | 0.02 |
| rs61914045 | A | G | 0.03 | 0.20 | 0.005 | 7.96E-09 | 154.82 | 6.55E-05 | TRUE | 0.29 |
| rs62535714 | A | G | 0.03 | 0.16 | 0.006 | 4.69E-09 | 157.60 | 6.83E-05 | TRUE | 0.20 |
| rs66511648 | C | T | 0.03 | 0.28 | 0.005 | 6.03E-10 | 179.50 | 7.65E-05 | TRUE | 0.21 |
| rs7152906 | C | T | 0.03 | 0.52 | 0.004 | 1.87E-09 | 166.28 | 7.20E-05 | TRUE | 0.08 |
| rs7241572 | A | G | 0.03 | 0.20 | 0.005 | 2.43E-09 | 169.97 | 7.15E-05 | TRUE | 0.72 |
| rs72948506 | A | G | 0.03 | 0.30 | 0.005 | 1.71E-08 | 146.87 | 6.36E-05 | TRUE | 0.69 |
| rs7538938 | C | T | 0.03 | 0.56 | 0.004 | 7.29E-09 | 155.35 | 6.81E-05 | TRUE | 0.31 |
| rs754287 | A | T | -0.03 | 0.37 | 0.005 | 1.31E-10 | 194.05 | 8.25E-05 | TRUE | 0.18 |
| rs7551758 | G | T | 0.03 | 0.53 | 0.004 | 5.11E-11 | 199.51 | 8.66E-05 | TRUE | 0.02 |
| rs76954012 | A | T | 0.04 | 0.09 | 0.007 | 2.41E-08 | 143.42 | 6.20E-05 | TRUE | 0.49 |
| rs7725715 | A | G | 0.03 | 0.53 | 0.004 | 1.61E-11 | 209.43 | 9.09E-05 | TRUE | 0.61 |
| rs843812 | A | G | 0.02 | 0.41 | 0.004 | 1.41E-08 | 149.07 | 6.35E-05 | TRUE | 0.03 |
| rs9364755 | G | A | 0.03 | 0.23 | 0.005 | 3.49E-08 | 140.28 | 6.16E-05 | TRUE | 0.72 |
| rs9529218 | T | C | -0.03 | 0.20 | 0.005 | 2.23E-10 | 187.24 | 7.92E-05 | TRUE | 0.36 |
| rs9536381 | T | C | 0.03 | 0.33 | 0.005 | 2.62E-08 | 142.95 | 6.14E-05 | TRUE | 0.26 |
| rs9831648 | T | G | -0.03 | 0.77 | 0.005 | 1.59E-08 | 149.30 | 6.30E-05 | TRUE | 0.33 |
| **Supplementary Table 4: Harmonised SNPs for the association between MDD and GlycA with exposure information** | | | | | | | | | | |

| Method | Description | |
| --- | --- | --- |
| Main analysis | | |
| Mendelian Randomisation (MR) | MR is an instrumental variable approach , using genetic variants as instruments for a modifiable exposure. It interrogates the causal effect of an exposure on an outcome, by utilising the random assortment of genetic variants (during gamete formation) from parents to offspring and as a result, the potential bias from confounding and reverse causation are minimised. MR is bound by three assumptions: (i) the genetic variants are statistically strongly associated with the exposure of interest and relevant to the population to which inference is being made (the relevance assumption); (ii) there is no confounding of the SNP-outcome association; and (iii) any effect of the genetic instrument on the outcome is only via the exposure [15] (supplementary figure 1). | |
| Inverse Variance Weighted (IVW) | A Wald ratio estimate is calculated for each genetic variant and summarised using the weighted regression of the SNP-exposure estimates on SNP-outcome estimates, where the intercept was constrained to zero. IVW assumes that there is no correlation between the association of the SNP exposure and SNP-pleiotropic path (the Instrument Strength Independent of Direct Effect (InSIDE) assumption) in the presence of horizontal pleiotropic paths. | |
| Methods to explore MR assumption | | |
| F-Statistic | Relevance assumption and weak instrument bias | Investigating the strength of genetic instruments using the F-statistics and proportion explained (R^2^) by each SNP. F-statistics >10 indicate that the estimates are not substantially biased by weak instruments [1] |
| MR-Egger | Horizontal pleiotropy. | Unlike the IVW, MR-Egger does not constrain the regression line to go through zero. Therefore, the MR-Egger regression represents the estimate of the causal effect controlling for unbalanced horizontal pleiotropy. A non-null MR-Egger intercept provides evidence of horizontal pleiotropy. MR-Egger has less statistical power than IVW and there should be no violation of the InSIDE assumption |
| Weighted Median | Horizontal pleiotropy | Provides an unbiased estimate when up to 50% of the SNPs used in the instrument violate the IV assumption |
| Weighted Mode | Horizontal pleiotropy | Uses the mode of the IVW empirical density function as the effect estimate |
| Cochran’s Q / Rucker’s Q statistic | Horizontal pleiotropy | Test for between-SNP heterogeneity in the IVW and MR-Egger analysis. |
| MR-Egger intercept | Horizontal pleiotropy | Under the assumption that pleiotropy is independent of the SNP-exposure association will give evidence for pleiotropic effects. |
| Sample definition/ underlying population | Confounding (via population stratification) | To mitigate bias caused by population stratification, which can confound the genetic instrument-outcome association and therefore violate the independence assumption, we restricted analyses to GWAS’ of the same underlying population (European ancestry participants only). |
| Radial MR | Horizontal pleiotropy | The method is similar to IVW but using a simulation-based approach fits a radial IVW model and provides an effect estimate, allowing outliers to be identified using Cochran’s Q statistic. The same can be done for a radial MR-Egger model. |
| Steiger filtering | Directionality of effects | Determines whether the proportion of variance explained by each SNP was larger in the exposure than that in the outcome. SNPs that did not pass Steiger filtering were excluded [2]. |
| MR Pleiotropy Residual Sum and Outlier global test (MR-PRESSO) | Horizontal pleiotropy | This method evaluates horizontal pleiotropy and corrects for it via outlier removal. It then tests for significant distortion in the causal estimate before and after outlier removal. |
| MR-LAP | Sample overlap, Winners Curse and weak instrument bias | MR-Lap uses cross-trait LD-score regression (LDSC) to simultaneously account and correct for winner’s curse, weak instrument bias and sample overlap [3]. |
| Supplementary Table 5: Description of MR, MR methods and sensitivity analyses | | |

|  | **Observed** | | |  | **Imputed (N=4021)** |
| --- | --- | --- | --- | --- | --- |
| **Variables** | **Categories** | **N** | **Mean (SE) for continuous variables Number (%) for categorical variables** | **% data imputed** | **Mean (SE) for continuous variables Number (%) for categorical variables** |
| Ethnicity | White | 3562 | 98.05 | 9.65 | 97.95 |
|  | Non-White | 71 | 1.95 |  | 2.05 |
| Sex | Male | 1696 | 42.18 | 0 | 42.18 |
|  | Female | 2325 | 57.82 |  | 57.82 |
| BMI at 18y clinic (kg/m^2^) |  | 3537 | 22.73 (0.07) | 12.04 | 22.74 (0.06) |
| BMI at 24y clinic (kg/m2) |  | 2996 | 24.74 (0.09) | 25.49 | 24.90 (0.09) |
| Age at 18y clinic in months |  | 3609 | 213.41 (0.08) | 10.25 | 213.39 (0.09) |
| Age at 24y clinic in months |  | 3025 | 293.17 (0.17) | 24.77 | 293.25 (0.18) |
| SMFQ score at 18y |  | 3392 | 6.45 (0.09) | 15.64 | 6.51 (0.09) |
| SMFQ score at 24y |  | 2718 | 6.84 (0.11) | 32.40 | 6.87 (0.11) |
| CIS-R at 18y | No depression diagnosis | 3052 | 92.54 | 17.98 | 92.26 |
|  | Diagnosis of depression | 247 | 7.46 |  | 7.74 |
| CIS-R at 24y | No diagnosis of depression | 2690 | 89.76 | 25.47 | 89.72 |
|  | Depression diagnosis | 307 | 10.24 |  | 10.28 |
| Average GlycA at age 18y (mmol/L) |  | 2918 | 1.22 (0.002) | 27.43 | 1.22 (0.002) |
| Average GlycA at age 24y (mmol/L) |  | 2836 | 1.23 (0.003) | 29.47 | 1.23 (0.003) |
| Maternal smoking during pregnancy | Never | 3104 | 84.44 | 8.58 | 84.37 |
|  | Temporary | 147 | 4.00 |  | 3.98 |
|  | Throughout | 425 | 11.56 |  | 11.65 |
| Maternal self-reported highest education qualification | Less than degree | 2808 | 79.19 | 11.81 | 79.50 |
|  | Degree or above | 738 | 20.81 |  | 20.50 |
| Smoking Status at 18y | Non-smoker | 1551 | 65.47 | 41.08 | 65.37 |
|  | Infrequent Smoker | 341 | 14.39 |  | 15.10 |
|  | Frequent Smoker | 477 | 20.14 |  | 19.53 |
| Smoking Status at 24y | Non-smoker | 1109 | 37.10 | 25.67 | 36.61 |
|  | Infrequent Smoker | 1382 | 46.24 |  | 46.15 |
|  | Frequent Smoker | 498 | 16.66 |  | 17.24 |
| Drinking Status at 18y | Non-Drinker | 544 | 15.77 | 14.20 | 16.12 |
|  | Infrequent Drinker | 1465 | 42.46 |  | 42.76 |
|  | Frequent Drinker | 1441 | 41.77 |  | 41.11 |
| Drinking Status at 24y | Non-Drinker | 50 | 2.22 | 43.87 | 2.44 |
|  | Infrequent Drinker | 1583 | 70.18 |  | 70.40 |
|  | Frequent Drinker | 623 | 27.60 |  | 27.16 |
| Household social economic position | Non-manual | 2915 | 86.55 | 16.24 | 85.96 |
|  | Manual | 453 | 13.45 |  | 14.04 |
| **Supplementary Table 6: The distributions of observed and imputed characteristics at ages 18y and 24y** | | | | | |

|  |  | **Model 1** | | **Model 2*** | |
| --- | --- | --- | --- | --- | --- |
|  | **N** | **Mean difference per SD increase in exposure** | **95% CI** | **Mean difference per SD increase in exposure** | **95% CI** |
| GlycA at 18y in relation to SMFQ score at 24y | 575 | 0.05 | -0.03, 0.13 | 0.01 | -0.08, 0.10 |
| SMFQ score at 18y in relation to GlycA levels at 24y | 575 | 0.04 | -0.05, 0.12 | 0.01 | -0.07, 0.09 |
|  | **N** | **OR** | **95% CI** | **OR** | **95% CI** |
| GlycA at 18y in relation to depressive episode at 24y | 575 | 1.28 | 0.96, 1.70 | 1.20 | 0.87, 1.66 |
| Depressive episode at 18y in relation to GlycA at 24y | **N** | **Mean difference per SD increase in exposure** | **95% CI** | **Mean difference per SD increase in exposure** | **95% CI** |
| Depression (as measured by CIS-R) at 18y on GlycA at 24y | 575 | 0.11 | -0.19, 0.41 | 0.02 | -0.17, 0.31 |
| **Supplementary Table 7: Bidirectional associations between continuous SMFQ and GlycA cross-sectionally at ages 18y and 24y using complete case analysis** * Adjusted for smoking status, drinking status, age in months at baseline, sex, ethnicity, maternal highest education qualification and BMI at 18y. | | | | | |

| **GWAS** | **Instrument** | **Outcome** | **Number of SNPs** | **Instrument F-statistics ^a^** | **Exposure**  **Sample Size** |
| --- | --- | --- | --- | --- | --- |
| Burges | GlycA | Depression | 51 | Min = 25.98  Median = 43.73  Max = 968.85 | 115,078 |
| Burges | GlycA | Depressive symptoms | 42 | Min=25.98  Median = 43.65  Max=968.85 | 115,078 |
| Howard *et al.* (2018)[4] | MDD | GlycA | 47 | Min = 137.83  Median = 160.00  Max =363.47 | 500,199 |
| Okbay *et al.* (2016) [5] | Depressive Symptoms | GlycA | 2 | Min = 47.83  Median = 49.07  Max =50.31 | 161,460 |
| Ahluwalia et al.  (2021) [6] | IL-6 | MDD | 1 | 2.71 | 3301 |
| Howard *et al.* (2018)[4] | MDD | IL-6 | 20 | Min = 164.98  Median= 139.48 Max=220.44 | 500,199 |
| **Supplementary Table 8*:* Genetic instruments and sample sizes used to estimate SNP-exposure and SNP-outcome associations**  ^a^Instrument strength F-statistics are based on the formulae $R^{2}=2*MAF*\left( 1-MAF \right)*{beta}^{2}$, where MAF=Minor allele frequency, and $F= \frac{R^{2}* ( N-2 )}{1-R^{2}}$ ; as described in Shim *et al.*^17^ and Palmer *et al.*^18^ previously.  These results are post-steiger filtering. | | | | | |

| **SNP** | **Effect Allele** | **Other Allele** | **beta** | **eaf** | **se** | ***p*** | **F** | **R^2^** | **steiger direction** | **steiger *p*** |
| --- | --- | --- | --- | --- | --- | --- | --- | --- | --- | --- |
| rs6684439 | T | C | 0.06 | 0.15 | 0.01 | 0.99 | 2.708 | 0.001 | False | 0.935 |
| **Supplementary Table 9: Harmonised SNPs for the association between IL-6 and MDD with exposure information** | | | | | | | | | | |

| SNP | Effect Allele | Other Allele | beta | eaf | se | pval | r2 | F.stat | *Steiger direction* | *Steiger p* |
| --- | --- | --- | --- | --- | --- | --- | --- | --- | --- | --- |
| rs1021363 | G | A | -0.03 | 0.6434 | 0.0045 | 2.29E-11 | 0.000413 | 206.6595 | TRUE | 0.651115 |
| rs10913112 | T | C | -0.0262 | 0.378 | 0.0045 | 4.53E-09 | 0.000323 | 161.5087 | TRUE | 0.662549 |
| rs12967143 | C | G | -0.0345 | 0.7012 | 0.0047 | 2.53E-13 | 0.000499 | 249.6022 | TRUE | 0.7218 |
| rs13037326 | T | C | 0.031 | 0.2597 | 0.0049 | 2.40E-10 | 0.00037 | 184.899 | FALSE | 0.552214 |
| rs1931388 | G | A | -0.0295 | 0.4042 | 0.0044 | 1.68E-11 | 0.000419 | 209.7461 | FALSE | 0.355174 |
| rs1950829 | G | A | -0.0297 | 0.5173 | 0.0043 | 4.74E-12 | 0.000441 | 220.4424 | FALSE | 0.721695 |
| rs2111592 | A | G | 0.0263 | 0.3141 | 0.0046 | 1.35E-08 | 0.000298 | 149.1217 | TRUE | 0.823155 |
| rs2214123 | G | A | -0.0261 | 0.6466 | 0.0045 | 8.56E-09 | 0.000311 | 155.7721 | TRUE | 0.662857 |
| rs2232423 | G | A | -0.062 | 0.1056 | 0.007 | 1.14E-18 | 0.000726 | 363.4675 | FALSE | 0.203539 |
| rs2418449 | C | T | -0.0281 | 0.281 | 0.0048 | 4.25E-09 | 0.000319 | 159.6458 | TRUE | 0.955458 |
| rs2522831 | C | T | 0.024 | 0.4739 | 0.0043 | 2.11E-08 | 0.000287 | 143.7055 | TRUE | 0.720365 |
| rs2568958 | A | G | 0.0382 | 0.6042 | 0.0044 | 2.90E-18 | 0.000698 | 349.3474 | TRUE | 0.911139 |
| rs2876520 | G | C | 0.026 | 0.4688 | 0.0043 | 2.24E-09 | 0.000337 | 168.465 | TRUE | 0.72582 |
| rs354155 | C | G | -0.0449 | 0.0923 | 0.0075 | 1.75E-09 | 0.000338 | 169.0264 | TRUE | 0.885307 |
| rs3807865 | A | G | 0.031 | 0.4105 | 0.0044 | 1.09E-12 | 0.000465 | 232.752 | FALSE | 0.461647 |
| rs4141983 | C | T | -0.0264 | 0.326 | 0.0046 | 9.69E-09 | 0.000306 | 153.2461 | TRUE | 0.969607 |
| rs4936276 | C | G | 0.0278 | 0.622 | 0.0044 | 3.57E-10 | 0.000363 | 181.8447 | FALSE | 0.832895 |
| rs7152906 | C | T | 0.0258 | 0.5196 | 0.0043 | 1.87E-09 | 0.000332 | 166.275 | TRUE | 0.709202 |
| rs7241572 | A | G | 0.0323 | 0.2047 | 0.0054 | 2.43E-09 | 0.00034 | 169.9701 | FALSE | 0.585933 |
| rs7551758 | G | T | 0.0283 | 0.5329 | 0.0043 | 5.11E-11 | 0.000399 | 199.5137 | FALSE | 0.720536 |
| rs843812 | A | G | 0.0248 | 0.4117 | 0.0044 | 1.41E-08 | 0.000298 | 149.0677 | FALSE | 0.390123 |
| rs9364755 | G | A | 0.0283 | 0.2262 | 0.0051 | 3.49E-08 | 0.00028 | 140.2772 | FALSE | 0.398088 |
| rs9536381 | T | C | 0.0255 | 0.3259 | 0.0046 | 2.62E-08 | 0.000286 | 142.95 | FALSE | 0.571334 |
| **Supplementary Table 10: Harmonised SNPs for the association between MDD and IL6 with exposure information** | | | | | | | | | | |

1.	Davies, N.M., M.V. Holmes, and G. Davey Smith, *Reading Mendelian randomisation studies: a guide, glossary, and checklist for clinicians.* BMJ, 2018. **362**: p. k601.

2.	Hemani, G., K. Tilling, and G. Davey Smith, *Orienting the causal relationship between imprecisely measured traits using GWAS summary data.* PLOS Genetics, 2017. **13**(11): p. e1007081.

3.	Mounier, N. and Z.J.b. Kutalik, *Correction for sample overlap, winner’s curse and weak instrument bias in two-sample Mendelian Randomization.* 2021.

4.	Howard, D.M., et al., *Genome-wide meta-analysis of depression identifies 102 independent variants and highlights the importance of the prefrontal brain regions.* Nat Neurosci, 2019. **22**(3): p. 343-352.

5.	Okbay, A., et al., *Genetic variants associated with subjective well-being, depressive symptoms, and neuroticism identified through genome-wide analyses.* 2016. **48**(6): p. 624-633.

6.	Ahluwalia, T.S., et al., *Genome-wide association study of circulating interleukin 6 levels identifies novel loci.* Hum Mol Genet, 2021. **30**(5): p. 393-409.
