## Supplementary figures for "Glycoprotein Acetyls and Depression: testing for directionality and potential causality using longitudinal data and Mendelian randomization analyses"

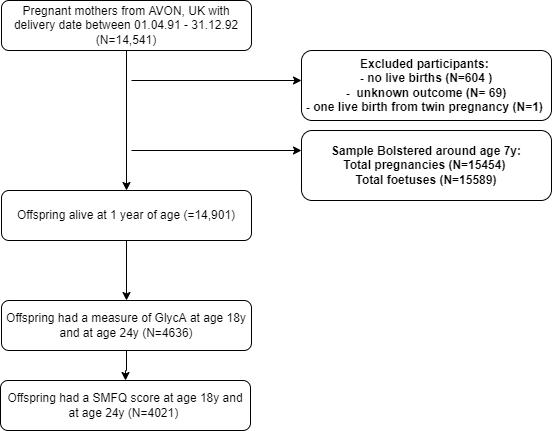

Figure 1: Flowchart of included ALSPAC participants

Figure 2: Forest Plot of IVW and sensitivity analyses showing the bidirectional relationship between genetically predicted IL-6 on MDD

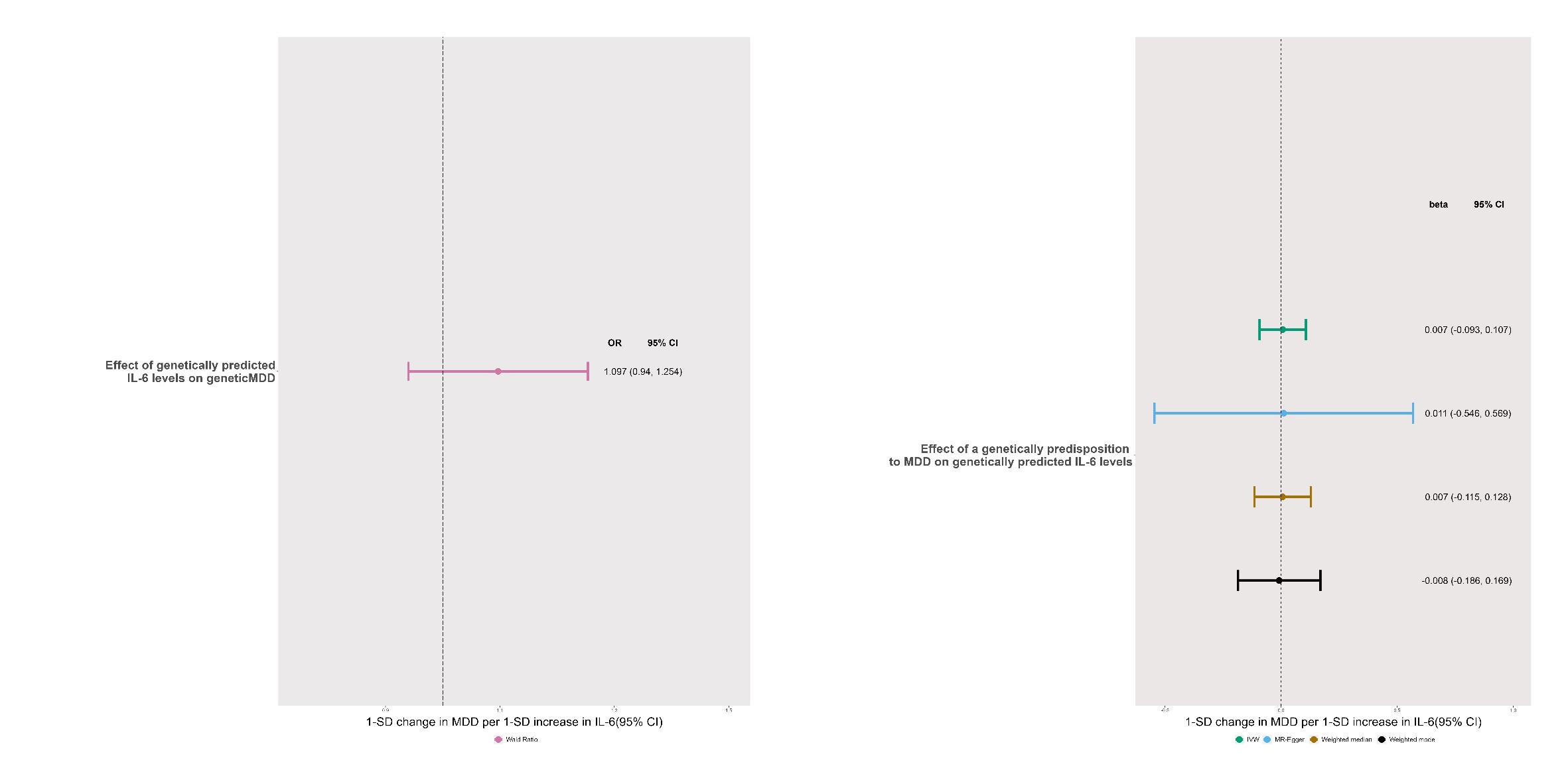

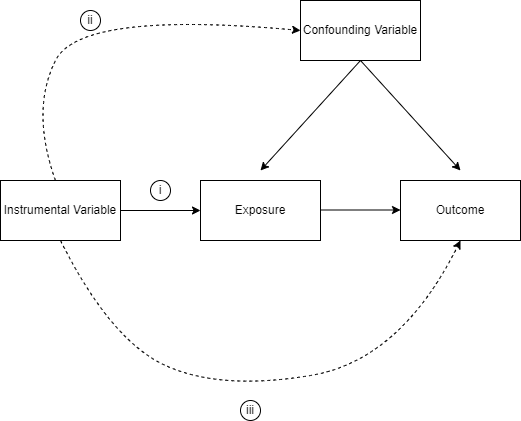

Figure 3: Directed Acyclic Graph (DAG) of MR and its assumption. (i) genetic variants associate with the exposure of interest; (ii) there is no confounding of the SNP-outcome association; and (iii) genetic variants exert effects on the outcome only via the exposure

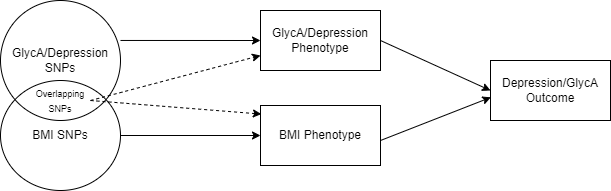

Figure 4: Directed Acyclic Graph demonstrating the direct (solid arrows) and indirect (dashed arrows) effects in the Multivariable Mendelian Randomisation analysis

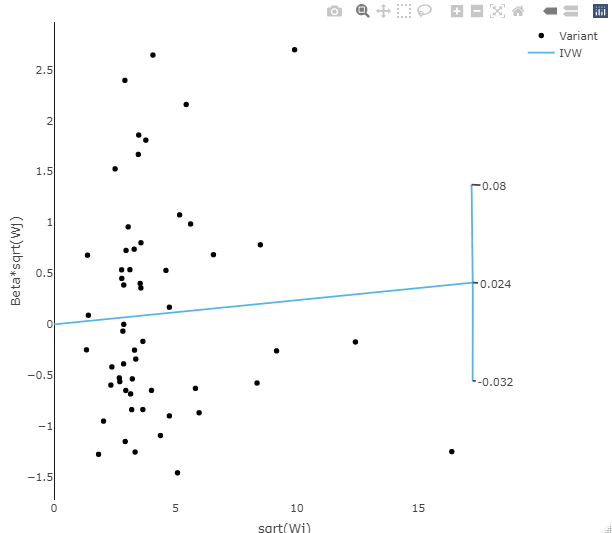

Figure 5: Radial IVW plots of SNP-GlycA versus SNP-depressive symptom level associations, with the IVW slope.

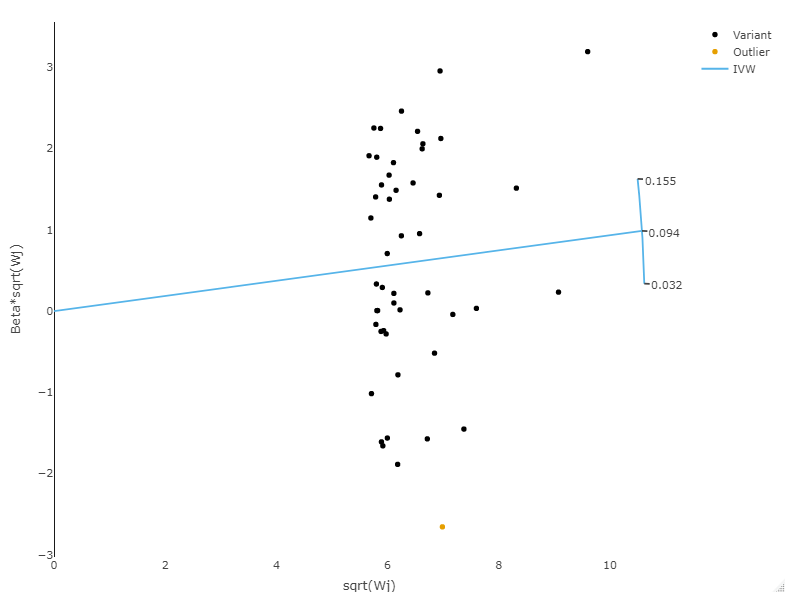

Figure 6: Radial IVW plots of SNP-MDD versus SNP-Glyca level associations, with the IVW slope.
